# Immunometabolic Blood Biomarkers of Developmental Trajectories of Depressive Symptoms: Findings from the ALSPAC Birth Cohort

**DOI:** 10.1101/2024.07.12.24310330

**Authors:** Ruby S. M. Tsang, Daniel Stow, Alex S. F. Kwong, Nicholas A. Donnelly, Holly Fraser, Inês Barroso, Peter A. Holmans, Michael J. Owen, Megan L. Wood, LINC Consortium, Marianne B. M. van den Bree, Nicholas J. Timpson, Golam M. Khandaker

## Abstract

Depression is associated with immunological and metabolic alterations, but immunometabolic characteristics of developmental trajectories of depressive symptoms remain unclear. Studies of longitudinal trends of depressive symptoms in young people could provide insight into aetiological mechanisms and heterogeneity behind depression, and origins of possible common cardiometabolic comorbidities for depression. Using depressive symptoms scores measured on 10 occasions between ages 10 and 25 years in the Avon Longitudinal Study of Parents and Children (n=7302), we identified four distinct trajectories: low-stable (70% of the sample), adolescent-limited (13%), adulthood-onset (10%) and adolescent-persistent (7%). We examined associations of these trajectories with: i) anthropometric, cardiometabolic and psychiatric phenotypes using multivariable regression (n=1709-3410); ii) 67 blood immunological proteins and 57 metabolomic features using empirical Bayes moderated linear models (n=2059 and n=2240 respectively); and iii) 28 blood cell counts and biochemical measures using multivariable regression (n=2256). Relative to the low-stable group, risk of depression and anxiety in adulthood was higher for all other groups, especially in the adolescent-persistent (RR_depression_=13.11, 95% CI 9.59-17.90; RR_GAD_=11.77, 95% CI 8.58-16.14) and adulthood-onset (RR_depression_=6.25, 95% CI 4.50-8.68; RR_GAD_=4.66, 95% CI 3.29-6.60) groups. The three depression-related trajectories vary in their immunometabolic profile, with evidence of little or no alterations in the adolescent-limited group. The adulthood-onset group shows widespread classical immunometabolic changes (e.g., increased immune cell counts and insulin resistance), while the adolescent-persistent group is characterised by higher BMI both in childhood and adulthood with few other immunometabolic changes. These findings point to distinct mechanisms and prevention opportunities for adverse cardiometabolic profile in different groups of young people with depression.

## Introduction

The first two decades of life represent a critical epoch for human neurodevelopment when most serious mental illnesses of adult life first emerge.^1^ Half of all lifetime cases of common mental disorders including depression and anxiety start by 14 years and 75% by 24 years.^2^ The first onset of clinically recognised depressive episodes typically occurs between the ages of 12 and 15 years^3^ and the increase in new onset of depression peaks between the ages of 15 and 18 years.^4^ Depressive symptoms in childhood and adolescence, including those below diagnostic thresholds, are associated with an elevated risk of depression and other psychiatric diagnoses subsequently in adulthood.^2, 5–7^ These findings highlight the need for studying depressive symptoms during early life.

Characterisation of longitudinal profiles of depressive symptoms during development could help understand the pathogenesis and heterogeneity of later depression, as different individuals may arrive at the same destination via different routes. There is growing evidence to suggest characteristic depression trajectories in childhood and adolescence are differentially associated with risk factors and outcomes. Existing studies have reported associations of a ‘high’ or ‘increasing’ depression trajectory with female sex, lower socioeconomic status, stressful life events, conduct issues, substance use, and parental psychopathology.^3, 8, 9^ Trajectories with higher symptom burden have been associated with subsequent depression and other psychiatric diagnoses, lower educational attainment, income and poorer psychosocial adjustment.^8–10^ However, less is known about underlying biological correlates of depression trajectories, including blood-based biomarker signatures. A better understanding of the biological correlates may help uncover mechanistic insights and identify accessible predictive markers for depression.

Existing literature suggests that depression and specific symptoms or symptom dimensions of depression are associated with immunometabolic dysfunction, but there is limited work on immunometabolic correlates of depression trajectories. Depression is associated with immunometabolic alterations such as chronic low-grade inflammation,^11, 12^ neuroendocrine dysregulations,^13^ as well as less favourable metabolic and lipid profiles.^12, 14^ Overall effect sizes for some of these associations are inconsistent, which could be partly due to clinical or phenotypic heterogeneity within cross-sectional studies.^12^ For instance, immunometabolic alterations appear to be more pronounced or common in individuals endorsing atypical energy-related symptoms of depression (e.g., hyperphagia, weight gain, hypersomnia, or leaden paralysis) as opposed to melancholic symptoms.^12, 15^ At the symptom level, inflammatory markers are particularly associated with somatic and neurovegetative symptoms of depression (e.g., fatigue, altered sleep and appetite) as opposed to psychological symptoms (e.g., hopelessness, excessive/inappropriate guilt).^16, 17^ Some of these findings are supported by Mendelian randomization analyses reporting a potentially causal link between inflammatory markers (e.g., C-reactive protein (CRP) or interleukin 6 (IL-6)) and fatigue, anhedonia, sleep problems, appetite and psychomotor changes.^18, 19^

The accumulation of risk model for chronic diseases posits that cumulative exposures across the life course result in diverging health trajectories and widening health inequalities as people age.^20^ By characterising depression trajectories, developmental windows when trajectories begin to diverge can be identified and we can then examine potential factors driving such divergence and biological dysregulations linked to subsequent disease risk. By studying the biomarker signatures of depression trajectories, we may also gain further insight into the origins of higher levels of cardiometabolic multimorbidity in individuals with depression.^21, 22^

The aims of the current study were threefold: (i) to model depressive symptom trajectories from childhood to early adulthood to classify individuals into more homogeneous subgroups, (ii) to examine associations between these subgroups and risk of psychiatric and cardiometabolic outcomes in early adulthood, and (iii) to examine associations of these subgroups with clinical and blood immunometabolic markers including proteomic, metabolomic and biochemical measures in early adulthood. By examining the broader biomarker signature across different domains including the immune proteome, metabolome, and clinical biochemistry, we aim to provide more comprehensive insights into biological pathways and systems possibly involved in the development and persistence of depressive symptoms in young people.

## Materials and Methods

### Description of cohort

This study uses data from the Avon Longitudinal Study of Parents and Children (ALSPAC). Pregnant women resident in the former county of Avon, United Kingdom (UK) with expected dates of delivery between 1^st^ April 1991 and 31^st^ December 1992 were invited to take part in the ALSPAC study. The initial recruitment enrolled 14541 pregnancies, which resulted in 14062 live births and 13988 infants still alive at 12 months. Further recruitment of eligible participants took place when the oldest children were approximately seven years of age; the total sample size for analyses using any data collected after the age of seven is therefore 15447 pregnancies; of these, 14901 children were alive at 12 months of age.^23, 24^

The study website contains details of all the data that is available through a fully searchable data dictionary and variable search tool: http://www.bristol.ac.uk/alspac/researchers/our-data/

### Data

#### Sociodemographic and health variables

Sociodemographic characteristics used to characterise the identified depressive symptom trajectories include sex, ethnicity, maternal education, maternal occupational social class, socioeconomic deprivation, and family adversity during pregnancy. Health characteristics examined include smoking, risky alcohol use, carotid intima-media thickness, carotid-femoral pulse wave velocity, metabolic syndrome and its components, obesity, and psychiatric outcomes and medications. Detailed description of these variables as well as those included as covariables in the biomarker analyses are presented in **Methods S1**.

#### Depressive symptoms

Self-reported depressive symptoms were assessed using the 13-item Short Mood and Feelings Questionnaire (SMFQ).^25^ We used data collected on 10 occasions between the ages of 10 and 25 years (ages 10, 12, 13, 16, 17, 18, 21, 22, 23, 25), ending with the last questionnaire administered in 2017-2018, prior to the start of the COVID-19 pandemic (see **Table S1**). Questions were answered based on the two-weeks prior to completing the questionnaire. Each SMFQ item is scored as 0 = “not true”, 1 = “sometimes true” and 2 = “always true”, resulting in a total SMFQ sum score 0-26 (higher score reflects more symptoms). For individuals who had missing data on fewer than three questions, score was imputed to the median value for missing items. For each time-point, those with missing data on more than three questions had their total score recoded as missing.

#### Circulating blood biomarkers

For this analysis, blood biomarkers were assayed in blood samples collected at the face-to-face research clinic undertaken at 24 years. A total of 92 circulating inflammatory proteins were measured using the Olink Target 96 Inflammation panel (Olink Analysis Service, Uppsala, Sweden); proteins with ≥50% values below the limit of detection (LOD) were excluded leaving 67 proteins to be included (**Table S2**). Over 220 metabolomic features (148 metabolites and 77 ratios) were quantified using a high-throughput ^1^H-NMR spectroscopy-based platform (Nightingale Health, Helsinki, Finland) using a standardised protocol and parameters described elsewhere.^26–28^ Lipoprotein subclasses were excluded from the analysis to minimise redundancy of information, leaving a subset of 57 metabolomic features (9 cholesterol measures, 12 apolipoproteins and lipids measures, 3 lipoprotein particle sizes, 16 fatty acids and saturation measures, 3 glycolysis-related metabolites, 8 amino acids, 3 ketone bodies, 2 fluid balance-related measures and 1 inflammation-related measure) to be included in the analysis (**Table S3**). All 26 blood count and chemistry measures collected at the same clinic interaction were included (**Table S4**). Additionally, we computed the aspartate aminotransferase/alanine aminotransferase (AST/ALT) ratio and the Homeostatic Model Assessment for Insulin Resistance (HOMA-IR). Further information on data collection and processing of all blood biomarkers are presented in **Methods S1**.

#### Covariables

Prior to statistical analysis, we plotted a directed acyclic graph (DAG) showing theoretical relationships between depressive symptom trajectories (independent variable), immunometabolic markers (dependent variables) and important covariables based on the literature (**Figure S1**). The minimum adjustment set of confounders included in models was sex at birth, maternal education, maternal occupational social class, and body mass index (BMI) at age 10.

### Statistical analysis

#### Characterisation of depressive symptom trajectories

Latent class trajectory modelling was performed using the *lcmm* R package^29^ to identify subgroups with distinct SMFQ trajectories. This type of modelling seeks to identify homogenous groups of individuals with similar trajectories within a heterogeneous population by combining a latent class model and a mixed model. Models are estimated within the maximum likelihood framework.^29^ The *lcmm* package distinguishes time of measurement and occasion, so individuals with missing data can still be included; we included those with at least three measurements for better modelling of non-linear trajectories. A multi-step approach adapted from the model selection framework suggested by Lennon et al.^30^ and van der Nest et al.^31^ was used, with the order of steps changed to address potential overextraction of latent classes from model under-specification as reported in the simulation literature^32, 33^. The steps followed:

1. Scope literature and inspect plots to inform polynomial order and potential number of classes. We modelled smfq ∼ age + age^2^ and estimated models up to six latent classes.
2. Estimate growth mixture models (GMM) with random intercepts and class-specific proportional random-effect variance-covariance matrix with increasing number of classes. Select the most appropriate number of classes *k* based on model convergence, model fit (Bayesian information criterion (BIC), Integrated Completed Likelihood (ICL), and relative entropy), smallest class size ≥5% and visual inspection of the trajectories.
3. Test alternative model structures with *k* classes – GMM with random intercepts and common random-effect variance-covariance matrix, and group-based trajectory models (GBTM); compare model fit indices, smallest class size and visually inspect trajectories as above, and assess model adequacy (average posterior probabilities ≥0.7 and odds of correct classification ≥5 for all classes).
4. Refine trajectory shape by testing up to second-degree fractional polynomials including (−2, −1, −0.5, 0, 0.5, 1, 2, 3) where 0 refers to log X and repeated polynomials refer to (X^i^ + X^i^ * log X). Select final model based on model convergence and model fit.

Age (in years) was used as the time variable in all models. No covariables were included in these latent class mixed models as the aim is to describe the trajectories; covariables were accounted for in the next step when testing for associations with phenotypes of interest and biomarkers. For each model, an automatic grid search with 50 sets of random initial values and up to 10 iterations was run to reduce the odds of the model converging towards a local maximum and then up to 500 iterations were allowed for the final estimation. Using the selected model, posterior probabilities for class membership were then estimated and individuals were assigned to the class of highest posterior probability in the entire sample using the *predictClass* function. We first performed the latent class mixed modelling on the subsample with three or more data points and then predicted class membership in the entire sample to reduce uncertainty in the modelling stage and to maximise sample sizes in the subsequent analyses. Additional information on the modelling is presented in **Supplementary Methods S1,** R scripts are provided in **Supplementary Methods S2**. The reporting of this study adheres to the Guidelines for Reporting on Latent Trajectory Studies (GRoLTS)^34^ (**Table S5**).

#### Associations with clinical and sociodemographic variables

Sociodemographic characteristics are stratified by trajectory class and summarised using mean (SD), median [interquartile range] or count (%) as appropriate, with differences between trajectories tested with chi-square or Kruskal-Wallis tests. Associations of trajectory membership with psychiatric or cardiometabolic outcomes of interest at age 24 or 28 years were tested using multivariable linear or logistic regressions, using the largest trajectory class as the reference group and adjusting for sex, maternal education, occupational social class and BMI at age 10. These variables are described in detailed in **Supplementary Methods S1**.

#### Associations with immunometabolic biomarkers

For both proteomic and metabolomic data, associations between depressive symptom trajectories and markers were evaluated using multiple linear models fitted in the *limma* R package.^35^ *Limma* uses an empirical Bayes method to moderate the standard errors of the estimated log-fold changes by borrowing strength from linear models of the other analytes and allowing for different variability between analytes and between samples. Planned contrasts of each of the intermediary trajectories against the trajectory with the most individuals were conducted. With the blood count and clinical chemistry data, linear regressions were fitted with blood markers as dependent variables and SMFQ trajectory class as the independent variable.

For each of these markers, the basic model included sex, maternal education and maternal occupational social class as covariables and the adjusted model further included BMI at age 10 as a covariable. Correction for multiple testing was performed for each set of models using the Benjamini-Hochberg procedure, using a false discovery rate (FDR) q-value threshold of <0.1. This threshold was chosen due to the large number of biomarkers tested, a relatively small sample size and the exploratory nature of this work. R scripts for these analyses are provided in **Supplementary Methods S2**.

#### Sensitivity analyses

To address potential error carried over from the probabilistic latent class assignment into the association analyses, we performed two sets of sensitivity analyses, the first set by restricting the sample to individuals who had a modal posterior probability ≥0.7, and the second set by using the individuals’ posterior probabilities for each latent class as separate terms in the models.

Data extraction and initial data cleaning was performed in StataMP version 17.^36^ Further data preparation and statistical analyses were conducted in R versions 4.1.1 and 4.2.1,^37^ using packages *tidyverse* (v2.0.0), *lcmm* (v2.0.2), *LCTMtools* (v0.1.3), *tableone* (version 0.13.2), *marginaleffects* (v0.25.0), *knitr* (v1.43), *kableExtra* (v1.3.4), *limma* (v3.54.2), and *broom* (v1.0.5). Plots were generated using *ggplot2* (v3.4.2), *ggpubr* (v0.6.0), and *ggrepel* (v0.9.3).

#### Ethical approval

Ethical approval for the ALSPAC study was obtained from the ALSPAC Ethics and Law Committee and the Local Research Ethics Committees. Consent for biological samples has been collected in accordance with the Human Tissue Act (2004). Informed consent for the use of data collected via questionnaires and clinics was obtained from participants following the recommendations of the ALSPAC Ethics and Law Committee at the time.

## Results

### Sample

Latent class trajectory modelling was performed on data from 7302 participants who had SMFQ scores available from at least three time-points between ages 10 and 25 years. Once the best-fitting model was identified, posterior probabilities and class membership were estimated in the entire sample, and 9595 individuals were assigned class membership. Of these 9595 individuals, 2256 had sufficient biomarker and complete covariable data to be included in the biomarker analyses (**Figure S2**).

### Depressive symptom trajectories from childhood to early adulthood

Following comparison of model fit and adequacy statistics and visual inspection of trajectory plots (**Table S6 and Figures S2-S3**), a four-class group-based trajectory model was identified as best describing the data. As shown in **Figure 1**, the four identified depressive symptom trajectories from childhood to early adulthood can be described as follows: low-stable – those who consistently had no or low levels of depressive symptoms (69.6%, n=6680), adolescent-limited – those who had elevated depressive symptoms in childhood/adolescence that decreased over time (13.3%, n=1280), adolescent-persistent – those who had elevated depressive symptoms in childhood/adolescence that remained high into adulthood (7.0%, n=672) and adulthood-onset –those who started with low levels of depressive symptoms that increased in late adolescence/early adulthood (10.0%, n=973).

**Figure 1.**
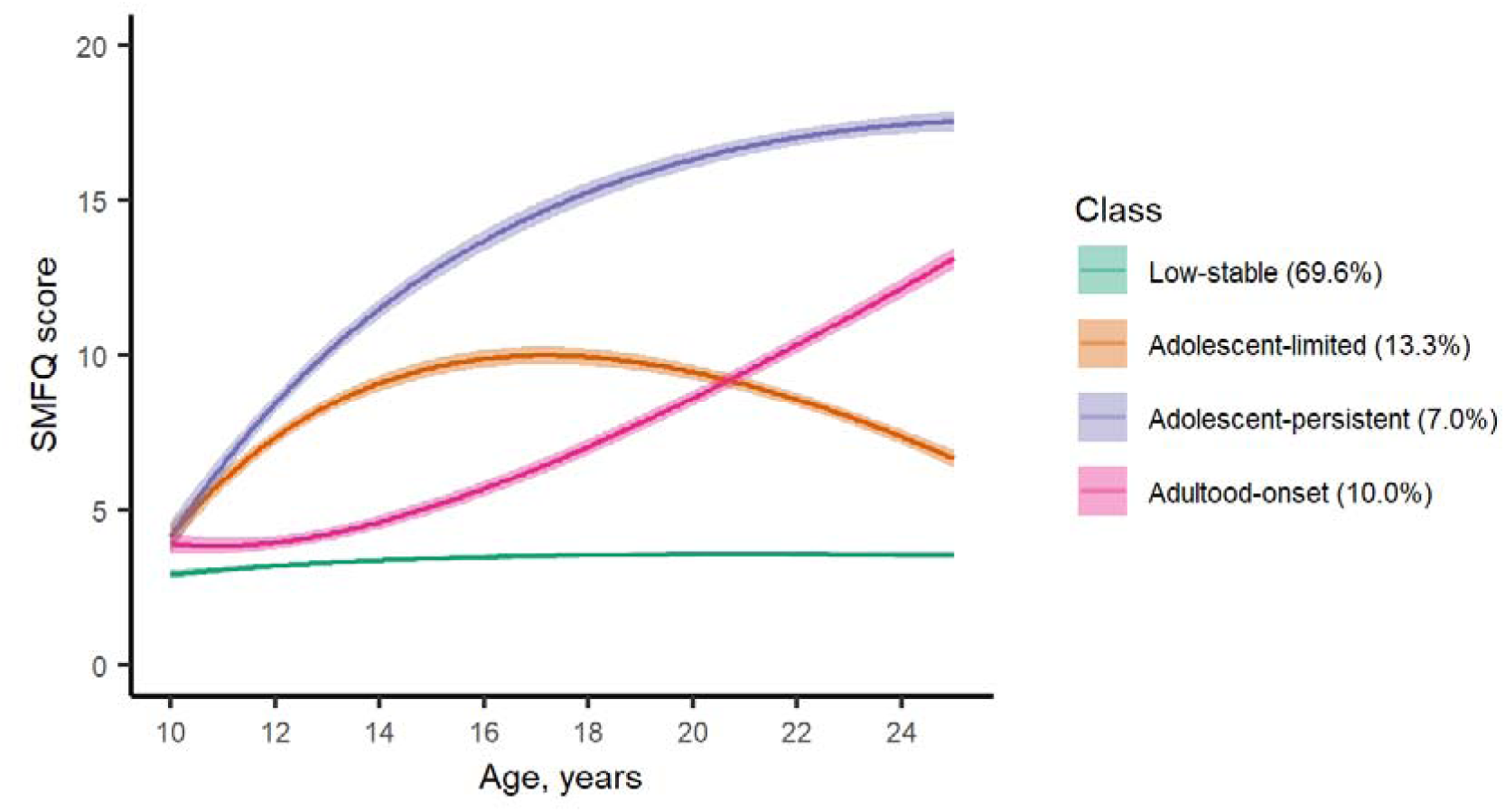
Predicted Marginal Mean Depressive Symptom Trajectories from Childhood to Early Adulthood in the ALSPAC Cohort.

### Characteristics of depressive symptom trajectories

Descriptive statistics for characteristics of these individuals, stratified by trajectory, are presented in **Table 1** below. There were more women in all three depression-related trajectories: adolescent-limited (66.3%), adolescent-persistent (76.5%), and adulthood-onset trajectories (64.6%). Additionally, the adolescent-persistent trajectory was associated with lower maternal education and greater family adversity during pregnancy. Descriptive statistics for the same characteristics of the subset of individuals who were included in the biomarker analyses are presented in **Table S7**.

**Table 1.**
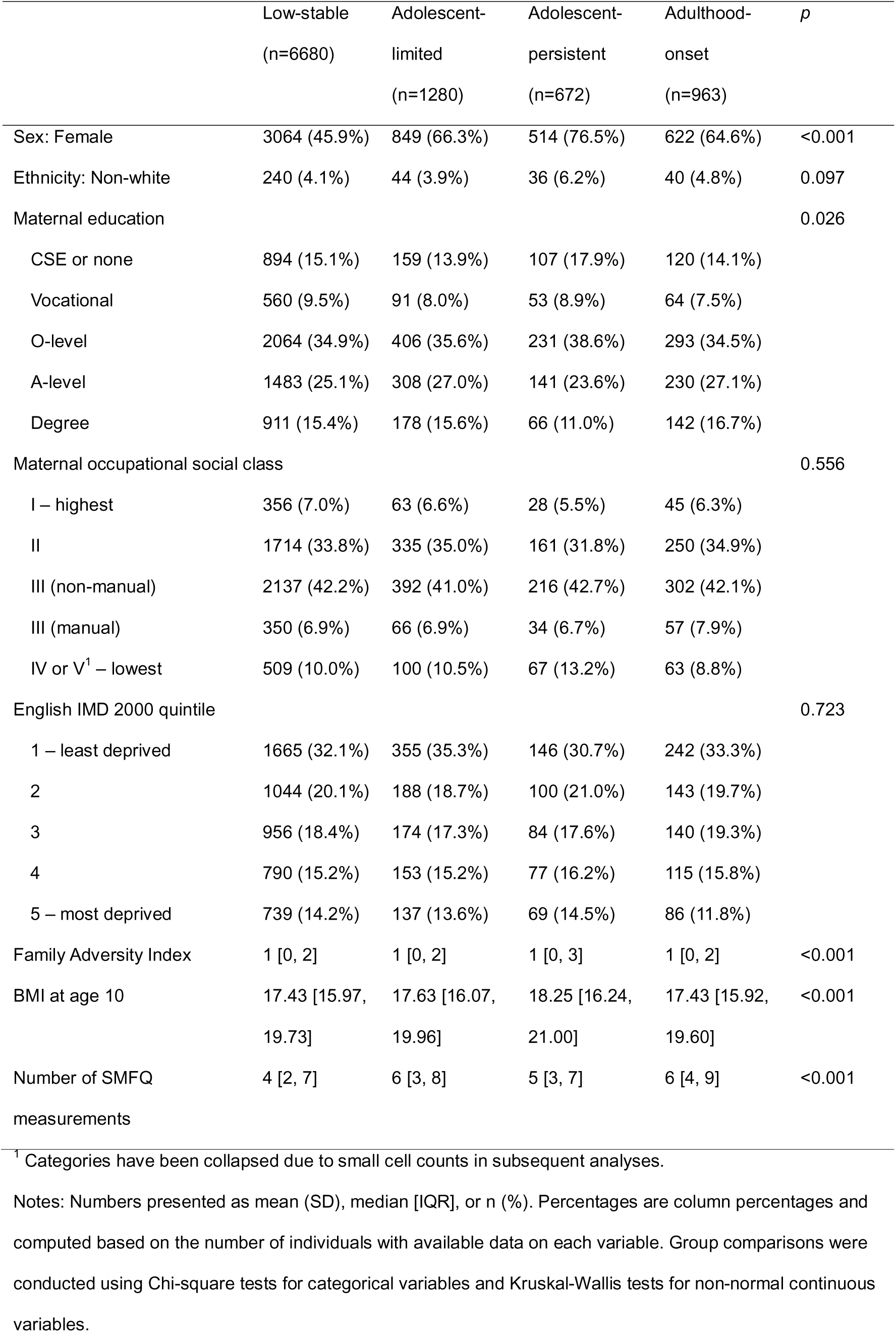
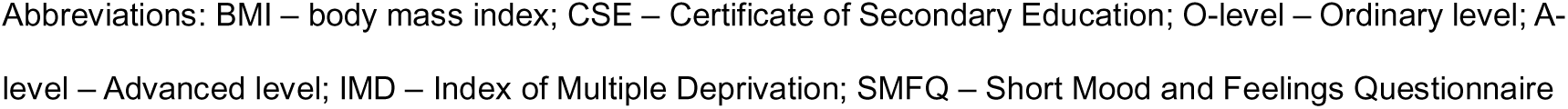
Characteristics of depressive symptom trajectories in the ALSPAC birth cohort.

|  | Low-stable<br>(n=6680) | Adolescent-limited<br>(n=1280) | Adolescent-persistent<br>(n=672) | Adulthood-onset<br>(n=963) | *p* |
| --- | --- | --- | --- | --- | --- |
| Sex: Female | 3064 (45.9%) | 849 (66.3%) | 514 (76.5%) | 622 (64.6%) | <0.001 |
| Ethnicity: Non-white | 240 (4.1%) | 44 (3.9%) | 36 (6.2%) | 40 (4.8%) | 0.097 |
| Maternal education |  |  |  |  | 0.026 |
| CSE or none | 894 (15.1%) | 159 (13.9%) | 107 (17.9%) | 120 (14.1%) |  |
| Vocational | 560 (9.5%) | 91 (8.0%) | 53 (8.9%) | 64 (7.5%) |  |
| O-level | 2064 (34.9%) | 406 (35.6%) | 231 (38.6%) | 293 (34.5%) |  |
| A-level | 1483 (25.1%) | 308 (27.0%) | 141 (23.6%) | 230 (27.1%) |  |
| Degree | 911 (15.4%) | 178 (15.6%) | 66 (11.0%) | 142 (16.7%) |  |
| Maternal occupational social class |  |  |  |  | 0.556 |
| I – highest | 356 (7.0%) | 63 (6.6%) | 28 (5.5%) | 45 (6.3%) |  |
| II | 1714 (33.8%) | 335 (35.0%) | 161 (31.8%) | 250 (34.9%) |  |
| III (non-manual) | 2137 (42.2%) | 392 (41.0%) | 216 (42.7%) | 302 (42.1%) |  |
| III (manual) | 350 (6.9%) | 66 (6.9%) | 34 (6.7%) | 57 (7.9%) |  |
| IV or V<sup>1</sup> – lowest | 509 (10.0%) | 100 (10.5%) | 67 (13.2%) | 63 (8.8%) |  |
| English IMD 2000 quintile |  |  |  |  | 0.723 |
| 1 – least deprived | 1665 (32.1%) | 355 (35.3%) | 146 (30.7%) | 242 (33.3%) |  |
| 2 | 1044 (20.1%) | 188 (18.7%) | 100 (21.0%) | 143 (19.7%) |  |
| 3 | 956 (18.4%) | 174 (17.3%) | 84 (17.6%) | 140 (19.3%) |  |
| 4 | 790 (15.2%) | 153 (15.2%) | 77 (16.2%) | 115 (15.8%) |  |
| 5 – most deprived | 739 (14.2%) | 137 (13.6%) | 69 (14.5%) | 86 (11.8%) |  |
| Family Adversity Index | 1 [0, 2] | 1 [0, 2] | 1 [0, 3] | 1 [0, 2] | <0.001 |
| BMI at age 10 | 17.43 [15.97, 19.73] | 17.63 [16.07, 19.96] | 18.25 [16.24, 21.00] | 17.43 [15.92, 19.60] | <0.001 |
| Number of SMFQ measurements | 4 [2, 7] | 6 [3, 8] | 5 [3, 7] | 6 [4, 9] | <0.001 |

<sup>1</sup> Categories have been collapsed due to small cell counts in subsequent analyses.
Notes: Numbers presented as mean (SD), median [IQR], or n (%). Percentages are column percentages and computed based on the number of individuals with available data on each variable. Group comparisons were conducted using Chi-square tests for categorical variables and Kruskal-Wallis tests for non-normal continuous variables.
Abbreviations: BMI – body mass index; CSE – Certificate of Secondary Education; O-level – Ordinary level; A- level – Advanced level; IMD – Index of Multiple Deprivation; SMFQ – Short Mood and Feelings Questionnaire

Lines showing predicted marginal mean depressive symptom trajectories with shaded areas representing 95% confidence intervals.

### Adulthood cardiometabolic and psychiatric outcomes associated with depressive symptom trajectories

Compared to the low-stable trajectory, after adjusting for sex, maternal education, maternal occupational class and BMI at age 10, all three depression-related trajectories were associated with ICD-10 diagnosis of depression, generalised anxiety disorder, being prescribed an antidepressant or anxiolytic at ages 24 and 28 years. However, the magnitude of association varied between the trajectories, with the risk for these outcomes being the highest for the adolescent-persistent trajectory (approximately 13-fold risk), followed by the adulthood-onset and adolescent-limited trajectories. The adolescent-persistent trajectory was additionally associated with obesity (**Table 2**). Unadjusted model results are presented in **Table S8**.

**Table 2.**
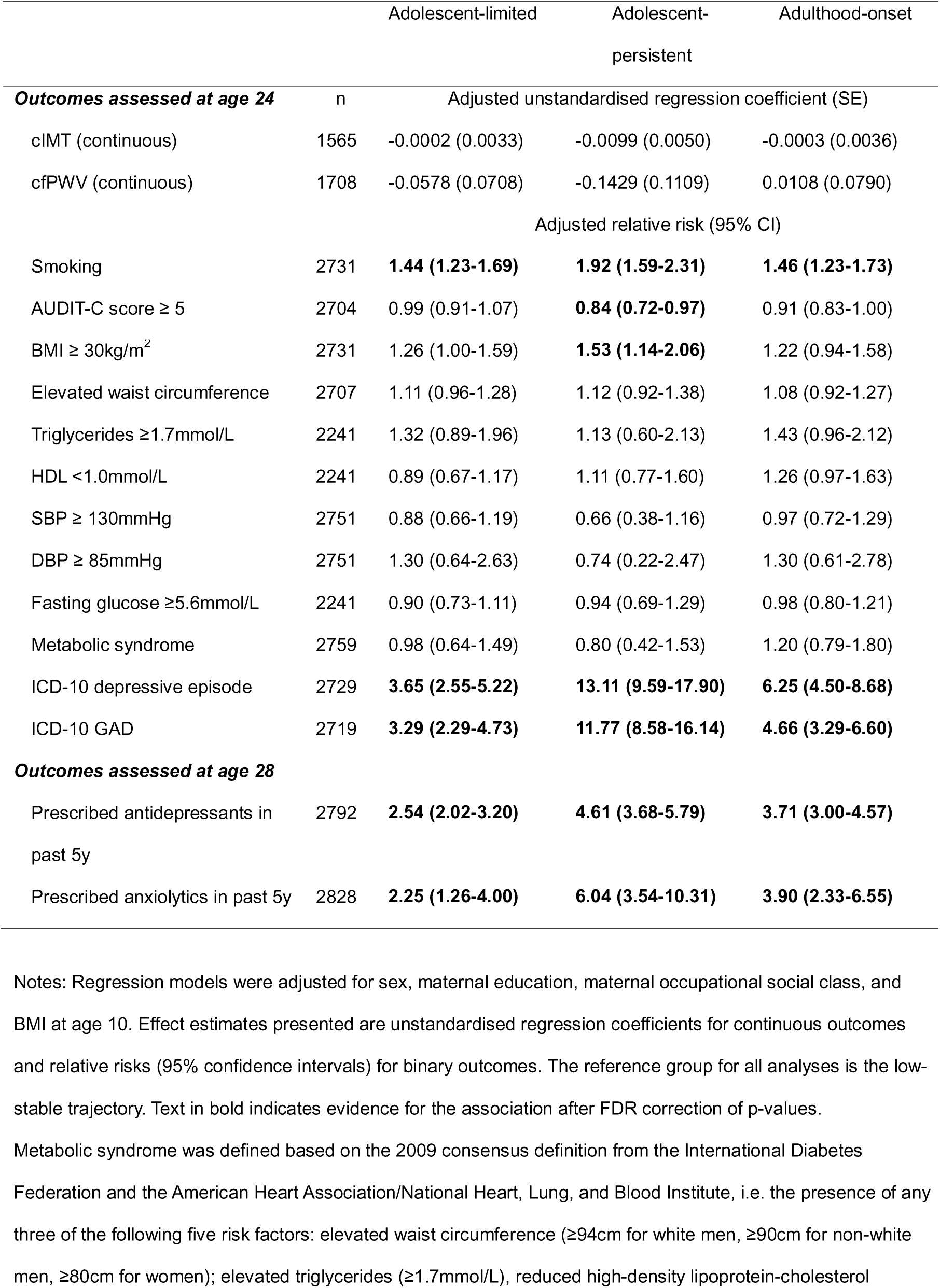
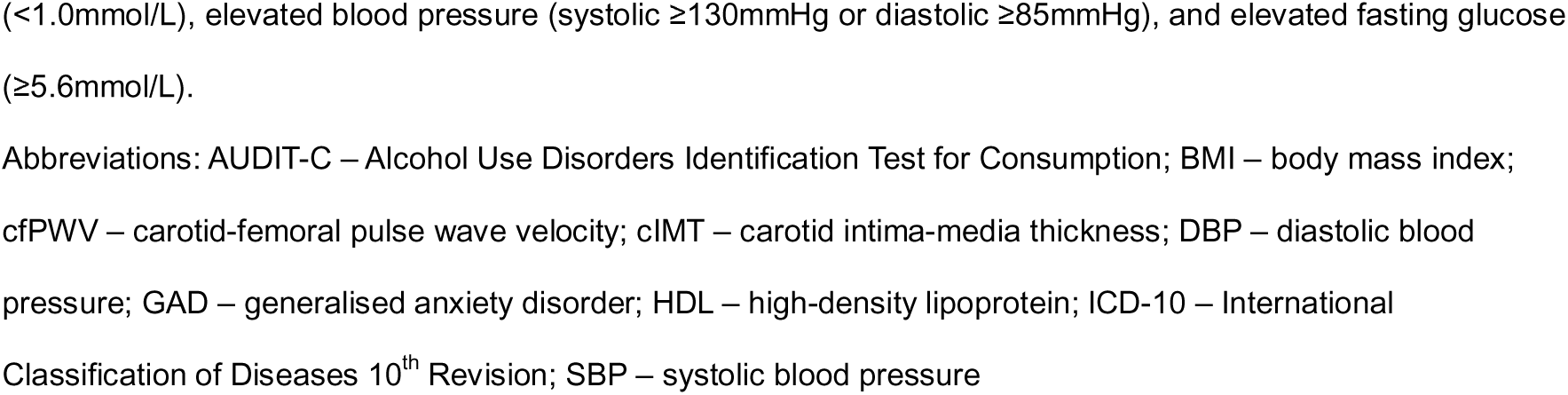
Associations of depressive symptom trajectories with anthropometric and cardiometabolic outcomes at 24 years and psychiatric outcomes at 24 and 28 years.

### Differentially abundant proteins associated with depressive symptom trajectories

Relative to the low-stable trajectory, after adjusting for sex, maternal education, maternal occupational class and BMI at age 10, one protein (C-C motif chemokine 25 [CCL25]) was upregulated in the adolescent-limited trajectory; four proteins (fibroblast growth factor 21 [FGF-21], hepatocyte growth factor [HGF], eukaryotic translation initiation factor [4E-BP1], and eotaxin-1 [CCL11]) were upregulated in the adolescent-persistent trajectory; and five proteins (FGF-21, fibroblast growth factor 19 [FGF-19], CUB domain-containing protein 1 [CDCP1], HGF, CCL11) were upregulated in the adulthood-onset trajectory (**Figure 2**). Full model results are presented in **Tables S9-10**.

**Figure 2.**
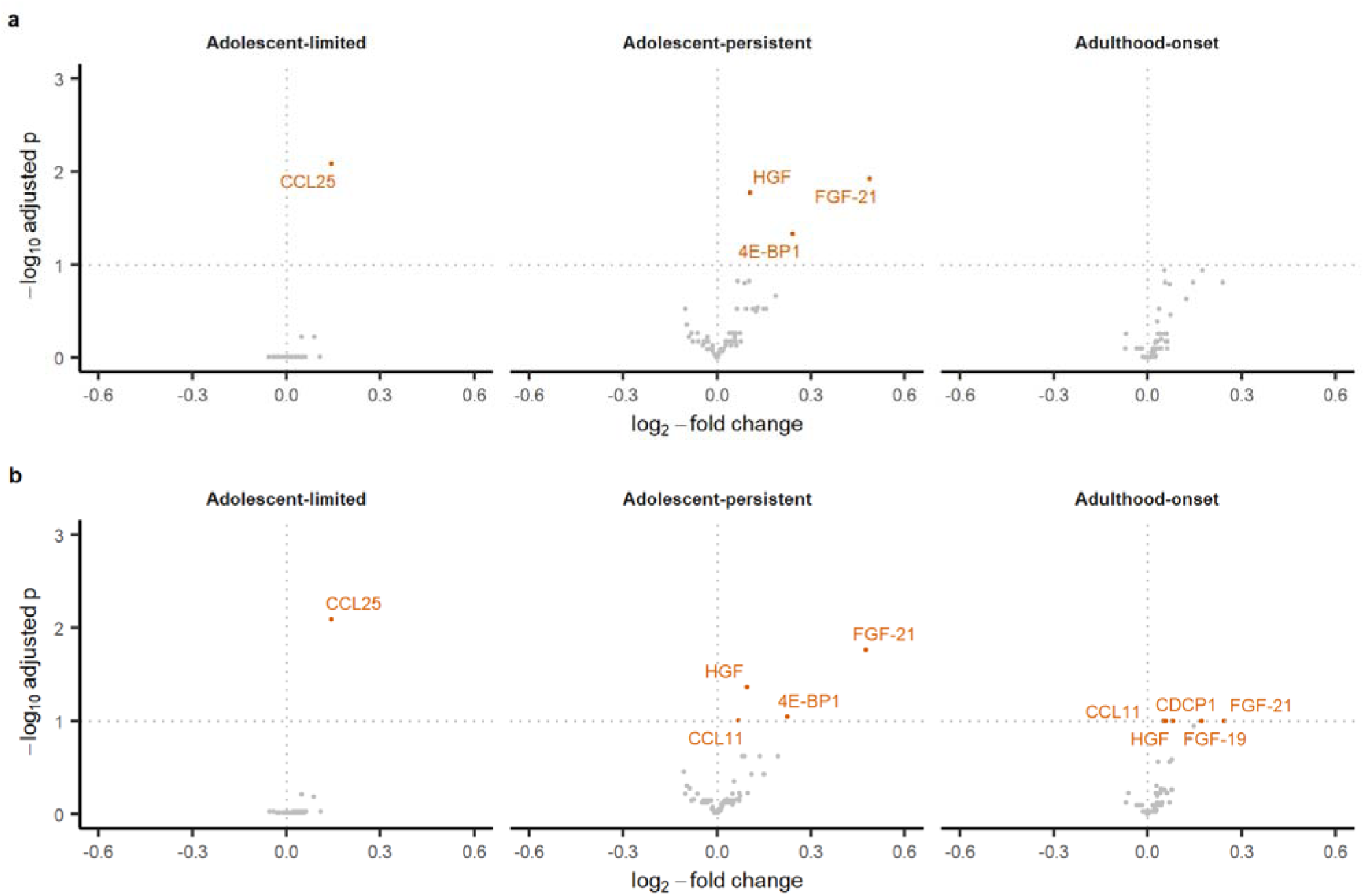
Volcano Plots Showing Differential Immune Protein Abundance Levels in Depressive Symptom Trajectories. Panels **a** and **b** show results from basic and adjusted models respectively. The reference group for all analyses is the low-stable trajectory. Abbreviations: 4E-BP1 – eukaryotic translation initiation factor; CCL11 – eotaxin-1; CCL25 – C-C motif chemokine 25; CDCP1 – CUB domain-containing protein 1; FGF-19 – oblast growth factor 19; FGF-21 – fibroblast growth factor 21; HGF – hepatic growth factor

### Differentially abundant metabolites associated with depressive symptom trajectories

Relative to the low-stable trajectory, after adjusting for sex, maternal education and occupational class, and BMI at age 10, creatinine was decreased in the adolescent-persistent trajectory. Three metabolite ratios (omega-3 to total fatty acids [omega-3/FA], docosahexaenoic acid to total fatty acids [DHA/FA], polyunsaturated fatty acids to total fatty acids [PUFA/FA] ratios) were decreased whereas the monosaturated to total fatty acids (MUFA/FA) and apolipoprotein B to apolipoprotein A1 (ApoB/ApoA1) ratios were increased in the adult-onset trajectory (**Figure 3**). Full model results are presented in **Tables S11-12**.

**Figure 3.**
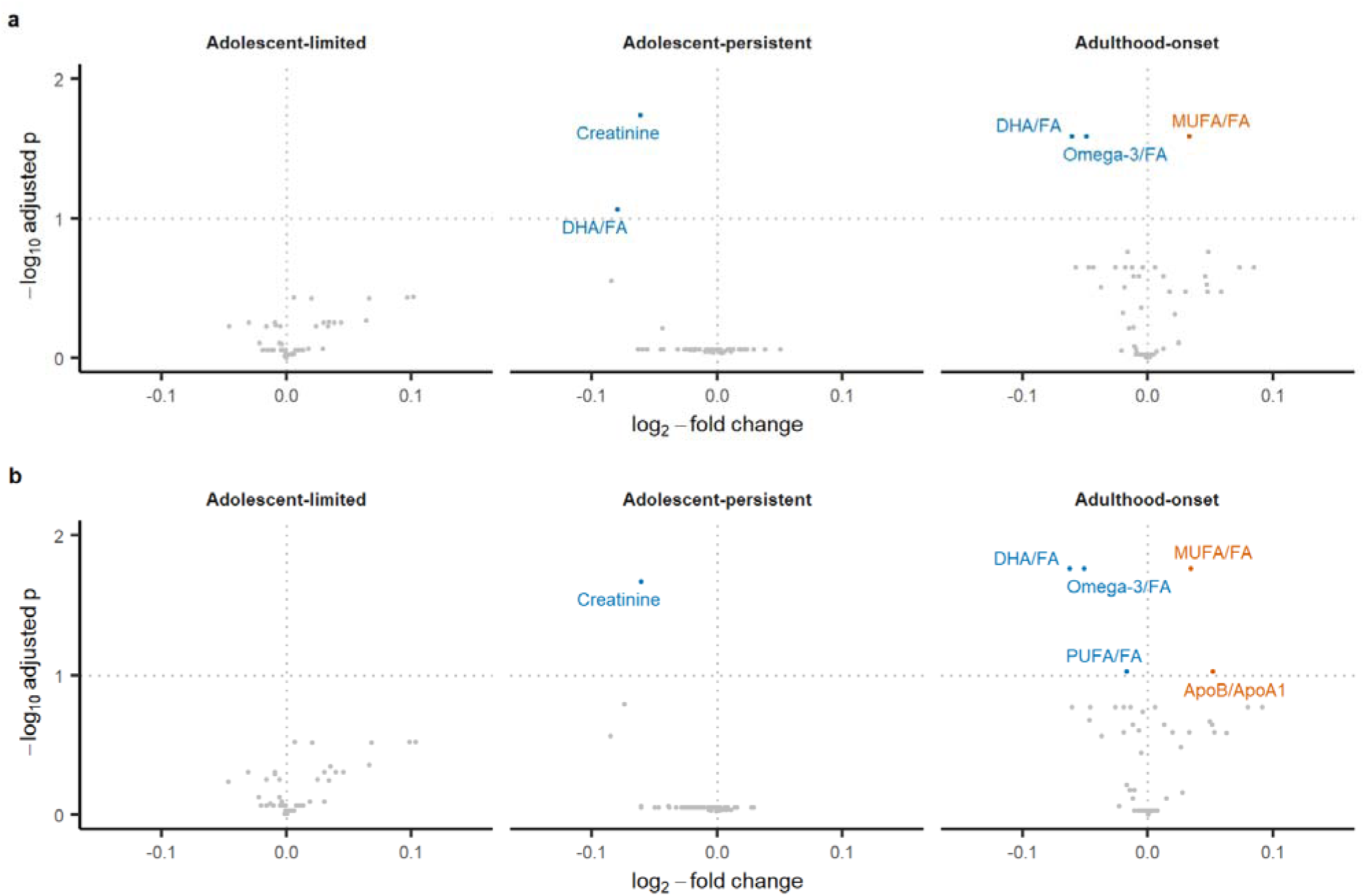
Volcano Plots Showing Differential Metabolite Abundance Levels in Depressive Symptom Trajectories. Panels **a** and **b** show results from basic and adjusted models respectively. The reference group for all analyses is the low-stable trajectory. Orange points indicate upregulation blue points indicate downregulation. Abbreviations: ApoB/ApoA1 – apolipoprotein B to apolipoprotein A1 ratio, DHA/FA – docosahexaenoic acid to total fatty acids ratio, MUFA/FA – monounsaturated fatty acids to l fatty acids ratio, Omega-3/FA – omega-3 fatty acids to total fatty acids ratio, PUFA/FA – polyunsaturated fatty acids/total fatty acids ratio

**Figure 4.**
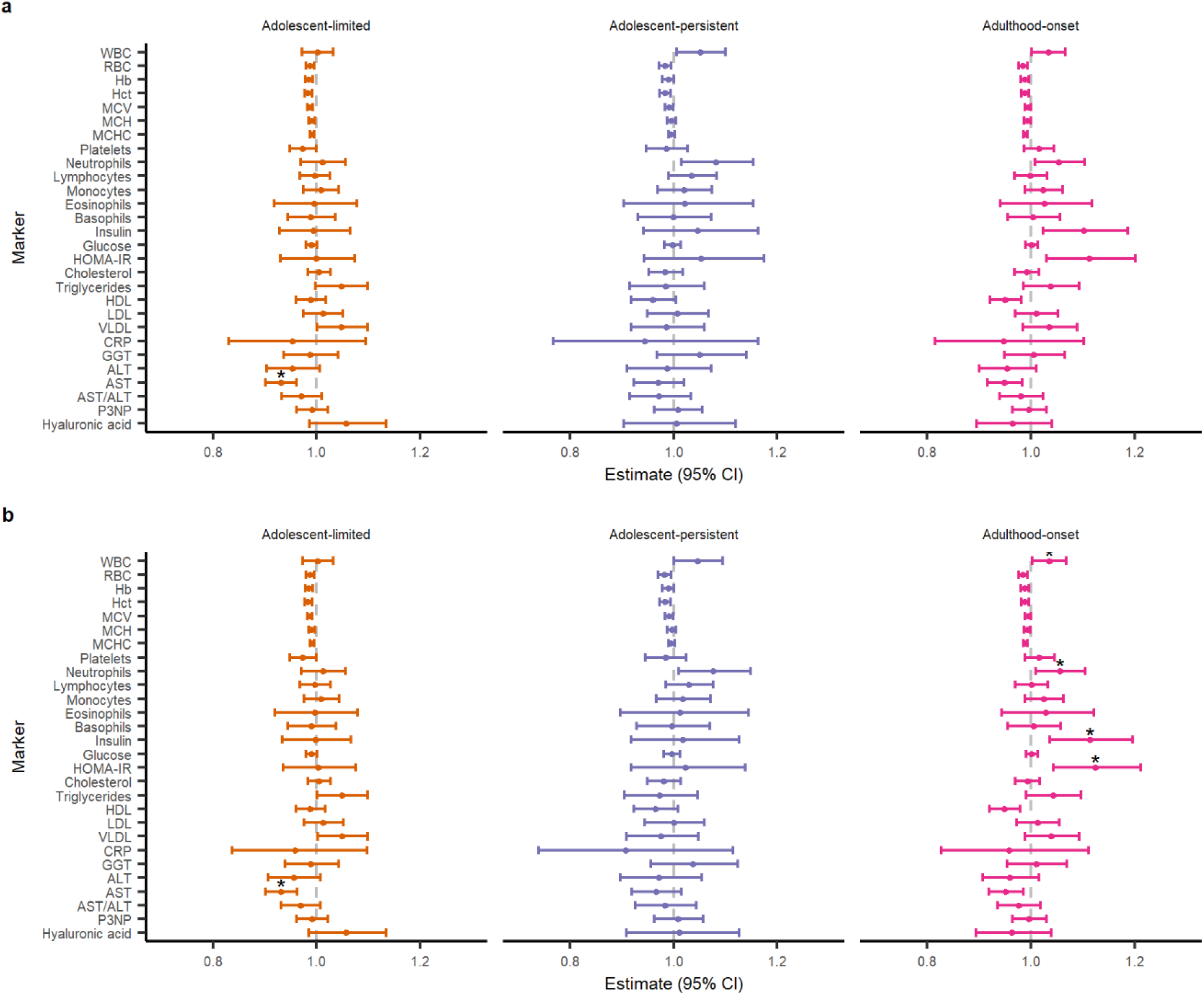
Differential Levels of Full Blood Count and Clinical Biochemistry Biomarkers in Different Depressive Symptom Trajectories. Dot-and-whisker plots showing effect estimates and 95% confidence intervals for each biomarker. The effect estimates and 95% confidence intervals from the models have been back-transformed into their original scale for ease of interpretation; effect estimates represent the percentage difference in mean levels of each biomarker for respective ectory, in relation to the low-stable trajectory (reference group). Panels a and b show results from basic and adjusted models respectively. Asterisks indicate evidence for the ociation after FDR correction of p-values. Abbreviations: WBC – white blood count, RBC – red blood count, Hb – haemoglobin, Hct – haematocrit, MCV – mean cell volume, MCH – mean cell haemoglobin, MCHC – an corpuscular haemoglobin concentration, HOMA-IR – Homeostatic Model Assessment for Insulin Resistance, HDL – high-density lipoprotein, LDL – low-density protein, VLDL – very low-density lipoprotein, CRP – C-reactive protein, GGT – gamma-glutamyl transpeptide, ALT – alanine aminotransferase, AST – aspartate notransferase, P3NP – procollagen-3 N-terminal peptide

### Blood count and clinical chemistry markers

Relative to the low-stable trajectory, after adjusting for sex, maternal education and occupational class, and BMI at age 10, there was evidence for decreased AST levels in the adolescent-limited trajectory, and increased HOMA-IR, insulin, neutrophil and white blood cell (WBC) counts in the adulthood-onset trajectory (Figure 5). Full model results are presented in **Tables S13-14**.

### Sensitivity analyses

The sensitivity analyses showed patterns of associations that are largely similar to those observed in the primary analyses, with consistent associations of depression-related trajectories with anthropometric, cardiometabolic and psychiatric outcomes, and top-ranking immunometabolic biomarkers with similar effect sizes in the same directions (**Supplementary Methods S3**).

## Discussion

Depression is a complex heterogeneous disorder, which poses a challenge for discovering biomarkers associated with disease onset and/or progression. We have taken a longitudinal approach to identifying blood-based biomarkers for depression by examining longitudinal patterns of depressive symptoms in the population during the critical developmental epoch of childhood, adolescence and early adulthood. Using data from a prospective birth cohort, we identified four longitudinal population subgroups based on repeated measures of depressive symptoms over a 15-year period from ages 10 to 25 years. We show that majority of participants (approximately 70%) have little or no depressive symptoms (low-stable group). We identified three depression-related groups which comprise a group with higher symptom levels during childhood and adolescence which later decrease (adolescent-limited group, 13%), a group with symptoms emerging during puberty that persist throughout adolescence through to adulthood (adolescent-persistent group, 7%), and a group with symptoms emerging during late adolescence/early adulthood and increasing thereafter (adulthood-onset, 10%). Earlier work examining latent depressive symptom trajectories in ALSPAC have reported varying numbers of latent trajectories, but all studies consistently identified a group with low/no symptoms, a group with symptoms emerging in early adolescence that persist, and a group with symptoms emerging in late adolescence/early adulthood.^10, 38, 39^ Differences in the number of trajectories identified may be due to sample inclusion/exclusion criteria, the number of repeated measures included, polynomials used to model the trajectories, as well as model selection criteria.

We examined health phenotypes and blood biomarkers associated with these subgroups for greater insight into the developmental course of depression and associated biomarkers. Our analyses show that compared to the low-stable group, risk of depression and anxiety in adulthood is higher for all three depression-related groups. However, such risk is particularly elevated risk for the adolescent-persistent group (11 to 13-fold risk) followed by the adulthood-onset group (four to six-fold risk). Interestingly, the group where higher levels of symptoms are mostly limited to adolescence, they still have a three-fold risk of depression in adulthood. This suggests that while their depressive symptoms do not persist at the same level as observed in adolescence, their symptoms do not fully return to the pre-morbid level either, and therefore are still more likely to meet criteria for a clinical diagnosis than those in the low-stable group. The literature also suggests that partial remission of depression is common and is a robust predictor for relapse.^40^

Having examined health phenotypic and blood proteomic, immunological and metabolic biomarker associations for these three groups, we show that the adolescent-limited group is distinct from the other two depression-related groups as it showed little immunometabolic alterations. In contrast, both the adolescent-persistent and adulthood-onset groups are associated with immunometabolic changes, but the exact pattern of associations varies between the two groups. The adolescent-persistent group was associated with higher BMI in childhood and adulthood, whereas the adulthood-onset group did not show this, but rather more widespread alterations in blood-based metabolic parameters including insulin resistance, insulin levels and changes in fatty acid ratios. Blood proteomic changes were largely similar between the two groups and involved proteins that mainly act as growth factors, cytokines and chemokines. While immunometabolic associations persisted given adjustment for childhood BMI in the adolescent-persistent group, the presence or absence of this BMI adjustment had an impact on some association estimates in the adolescent-persistent and adulthood-onset groups. While this may relate to the apparent differences in BMI by trajectories, it is difficult to distinguish this as an artefact of adjustment or a true impact of BMI.

Epidemiological studies consistently report a bidirectional relationship (both cross-sectionally and longitudinally) between obesity and depression,^41^ whereas Mendelian randomization studies support a causal role of BMI on major depressive disorder and depressive symptoms but not vice versa.^42, 43^ The comorbidity between obesity and depression is generally associated with poorer prognosis, with studies reporting associations with a more chronic course of depression in adulthood^44, 45^ as well as poorer treatment response.^46^ Our findings add to this evidence by showing an association between higher childhood BMI and persistent depressive symptoms between adolescence and early adulthood.

Many of the alterations observed in the adulthood-onset trajectory are already well studied markers of cardiometabolic disease risk. The ApoB/ApoA1 ratio is associated with cardiovascular diseases and metabolic syndrome,^47, 48^ and can be used to predict longer-term cardiovascular risk when measured in early life.^49, 50^ Higher values of HOMA-IR are associated with an increased risk of developing type 2 diabetes mellitus (T2DM), systemic arterial hypertension and non-fatal major adverse cardiovascular events.^51^ Lower blood omega-3 fatty acid concentrations are associated with poorer cardiovascular outcomes^52, 53^ and may also contribute to chronic systemic inflammation, whereas changes in MUFA, PUFA and DHA concentrations in early adulthood were associated with incident obesity, insulin resistance and elevated blood pressure 10 years later.^54^ Since dietary intake and supplementation are the main predictors of blood levels of omega-3 fatty acids,^55, 56^ with other lifestyle-related factors such as BMI, smoking and alcohol consumption also playing a role,^57^ this suggests that the adulthood-onset group may benefit from primary prevention strategies such as lifestyle modifications to prevent future cardiometabolic disease. Given that it is already well established that depression predisposes young people to accelerated atherosclerosis and early cardiovascular disease,^58^ further studies investigating the use of longitudinal course of depressive symptoms for risk stratification and prevention of cardiometabolic disease are warranted.

The overlapping proteomic signals observed between the adolescent-persistent and adulthood-onset trajectories potentially suggest shared underlying mechanisms (genetic or environmental) or shared biological responses to depression, which warrant further study for their roles in the pathophysiology of depression and cardiometabolic disease. FGF-21 is a novel regulator of glucose and lipid metabolism that mainly acts through an FGF receptor 1 (FGFR1)/β-klotho receptor complex and the Ras/Raf MAPK signalling pathway, which have been implicated in the pathophysiology of depression and therapeutic effects of antidepressants.^59–61^ Elevated circulating FGF-21 concentrations have also been shown to be associated with a range of cardiometabolic markers and diseases.^62–66^ HGF mediates inflammatory responses to tissue injury and regulates cell growth and morphogenesis through the activation of the

HGF/mesenchymal-epithelial transition factor (c-Met) signalling pathway, which has downstream effects on the Raf/MAPK and PI3K/Akt pathways. Altered HGF/c-Met signalling has been suggested to play a role in the pathogenesis of depression in adolescents through disrupting interneuron development.^67^ CCL11 is a chemokine involved in the selective recruitment of eosinophils into sites of inflammation and has been implicated in various allergic and inflammatory conditions.^68^ It can be transported across the blood-brain barrier^69^ and is an age-related systemic factor associated with reduced synaptic plasticity and impaired hippocampal-dependent learning and memory in mice.^70^ In humans, CCL11 levels increase with age,^70^ and there is emerging evidence to suggest that CCL11 levels are associated with psychiatric disorders.^71–75^ In summary, the overlapping proteomic signals between the adolescent-persistent and adulthood-onset trajectories highlight potential roles of physiological stress from lifestyle or environmental factors, disruptions in neurodevelopment and neurogenesis, and cellular senescence in the underlying vulnerability or biological response to depression, and may be key biomarkers relevant to illness pathogenesis.

An advantage of our work is that by examining depressive symptoms longitudinally using a latent class trajectory approach, we can account for population heterogeneity and obtain better characterisation of subgroups and their changes over time. Existing literature shows that depression is associated with alterations in various immunometabolic biomarkers, including increased inflammatory cytokines,^76, 77^ WBC,^78^ neutrophils,^78^ T-lymphocytes and other immune cell counts,^78^ HOMA-IR,^79^ insulin,^79^ lipids and fatty acids.^80, 81^ Using longitudinal data from young people, we add to this evidence base by showing that classical immunometabolic changes are particularly associated with an adulthood-onset trajectory, rather than other developmental subgroups of depressive symptoms, including one with persistent symptoms since adolescence. While developmental stages like childhood, adolescence and adulthood are in part socially constructed, our results show that the timings of divergence in depressive symptom trajectories are approximately aligned with the transitions between developmental stages when some of the major biological and environmental changes also take place (e.g., puberty, starting secondary school, and finishing school).

This study has several limitations. Firstly, the approach of treating assigned class membership as discrete in assessing relationships with other variables has been shown to underestimate the strength of the relationships.^82^ However, as we are mainly interested in subpopulations with different depressive symptom trajectories, this approach allows for easier interpretation and translation. Secondly, depression is episodic in nature and the use of polynomials cannot fully capture the dynamics of depressive symptom severity over time; however, this approach was chosen over other methods (e.g., splines) for model parsimony.

Furthermore, the sample size in this study is relatively small and may be underpowered to detect differences after stratifying individuals into four separate trajectories. For this reason, we did not further stratify our analyses by sex or other potentially relevant variables (e.g., BMI). While we have adjusted our biomarker analyses for several potential confounders, residual confounding could still be an issue. For instance, we have not accounted for medication use or chronic disease, but these are likely to be uncommon in young people. As the biomarkers were measured at age 24, which is after the onset of depressive symptoms in many individuals, further research is required to assess the direction and causality of associations we have identified.

In conclusion, we identified distinct developmental trajectories of depression from childhood to early adulthood, which show differential associations with cardiometabolic and psychiatric outcomes, and are characterised by distinct immunometabolic profiles. In particular, individuals with persistent depressive symptoms from childhood through to early adulthood were more likely to have higher BMI both in childhood and in early adulthood and few other immunometabolic changes, whereas individuals who develop depressive symptoms towards early adulthood show classical immunometabolic alterations in immune cell counts, insulin resistance and fatty acid profiles. These findings point to distinct mechanisms and prevention opportunities for adverse cardiometabolic profile in different groups of young people with depressive symptoms.

## Data Availability

ALSPAC data access is through a system of managed open access. Information on how to request access to ALSPAC data can be found on the ALSPAC website (http://www.bristol.ac.uk/alspac/researchers/access/).

## Acknowledgements

We are extremely grateful to all the families who took part in this study, the midwives for their help in recruiting them, and the whole ALSPAC team, which includes interviewers, computer and laboratory technicians, clerical workers, research scientists, volunteers, managers, receptionists and nurses. We would also like to thank members of the LINC study public advisory group for their contribution.

This work was funded by the Tackling Multimorbidity at Scale Strategic Priorities Fund programme (MR/W014416/1) delivered by the UK Medical Research Council (MRC) and the UK National Institute for Health Research (NIHR) in partnership with the UK Economic and Social Research Council and in collaboration with the UK Engineering and Physical Sciences Research Council. RSMT, DS and MLW are supported by this grant. ASFK is supported by a Wellcome Early Career Award (227063/Z/23/Z). NAD was supported by an NIHR Clinical Lectureship in General Adult Psychiatry. MBMvdB acknowledges additional funding support from the MRC (MR/W028395/1, MR/W020297/1, MR/T033045/1 and MR/S037667/1), NIMH (U01MH119758), the Wellcome Trust (226709/Z/22/Z) and Welsh Government (HS 22 04). NJT is a director of the MRC/ESRC/UKRI supported Population Research UK Coordination Hub (ES/Y008340/1), supported by a Wellcome Trust Investigator award (202802/Z/16/Z), is the PI of the Avon Longitudinal Study of Parents and Children (MRC & WT 217065/Z/19/Z), is supported by the University of Bristol NIHR Biomedical Research Centre (BRC-1215-2001), the MRC Integrative Epidemiology Unit (MC_UU_00011/1), CRUK and works within the CRUK Integrative Cancer Epidemiology Programme (C18281/A29019) and with support from CRUK (PRCPJT-May22\100028). GMK acknowledges funding support from the MRC (MC_UU_00032/6), which forms part of the MRC Integrative Epidemiology Unit at the University of Bristol. This grant also supports HF. GMK acknowledges additional funding from the Wellcome Trust (201486/Z/16/Z and 201486/B/16/Z), the MRC (MR/W014416/1; MR/S037675/1; and MR/Z50354X/1), and the NIHR Bristol Biomedical Research Centre (NIHR 203315). The views expressed are those of the authors and not necessarily those of the UK NIHR or the Department of Health and Social Care. The UK MRC and Wellcome (217065/Z/19/Z) and the University of Bristol provide core support for ALSPAC. This publication is the work of the authors and RSMT, NJT and GMK will serve as guarantors for the contents of this paper. A comprehensive list of grants funding is available on the ALSPAC website (http://www.bristol.ac.uk/alspac/external/documents/grant-acknowledgements.pdf).

## LINC Consortium members

Marianne B. M. van den Bree, George Kirov, Michael J. Owen, James T. R. Walters, Peter A. Holmans, Jane Lynch, Ioanna K. Katzourou, Nabila Ali, Lowri O’Donovan (Cardiff University, UK)

David A. van Heel, Sarah Finer, Daniel Stow (Queen Mary University of London, UK)

Golam M. Khandaker, Nicholas J. Timpson, John A. A. Macleod, Julie P. Clayton, Ruby S. M. Tsang, Jane Sprackman, Shahid Khan (University of Bristol, UK)

Inês Barroso, Rupert A. Payne (University of Exeter, UK)

Mark Mon-Williams, Megan L. Wood (University of Leeds, UK)

Hilary C. Martin (Wellcome Sanger Institute, UK)

Thomas Werge, Andrés Ingason, Morteza Vaez, Lam O. Huang (Institute of Biological Psychiatry, Denmark)

