## Supplementary Information for "Immunometabolic Blood Biomarkers of Developmental Trajectories of Depressive Symptoms: Findings from the ALSPAC Birth Cohort"

Methods S1. Additional materials and methods

Methods S2. R scripts for the latent class trajectory modelling and biomarker analyses

Methods S3. Sensitivity analyses

Figure S1. Directed acyclic graph for confounder assessment

Figure S2. Flow diagram illustrating sample selection for the latent class trajectory modelling and the biomarker analyses

Figure S3. Plots of predicted trajectories for models tested

Figure S4. Plots of predicted trajectories from the final model alongside individual trajectories

Table S1. Details of SMFQ time points included in the trajectory modelling

Table S2. List of 67 analysed inflammation proteomic markers

Table S3. List of 71 analysed metabolomic markers

Table S4. List of 28 analysed blood count and biochemistry markers

Table S5. Guidelines for Reporting on Latent Trajectory Studies (GRoLTS) checklist

Table S6. Summary of model fit and adequacy indices of the latent class mixed models fitted

Table S7. Characteristics of the ALSPAC subsample included in the biomarkers analyses, stratified by depression trajectory class

Table S8. Associations of depression trajectories with cardiometabolic outcomes at 24 years and psychiatric outcomes at 24 and 28 years – unadjusted model

Table S9. Results from the inflammation proteomics limma models – basic model

Table S10. Results from the inflammation proteomics limma models – adjusted model

Table S11. Results from the NMR metabolomics limma models – basic model

Table S12. Results from the NMR metabolomics limma models – adjusted model

Table S13. Results from the blood count and biochemistry linear regression models – basic model

Table S14. Results from the blood count and biochemistry linear regression models – adjusted model

References

**Methods S1 Additional Materials and Methods**

*Data*

Sociodemographic and clinical variables

1) Maternal education and maternal occupational social class

Maternal education and maternal occupational social class were collected at 32 weeks gestation and used as household socioeconomic status indicators. Maternal education was categorised into: Certificate of Secondary Education or no qualifications, vocational, O-level, A-level and university degree. Maternal occupational social class was categorised using the UK Registrar General’s occupational coding system: I professional, managerial and technical, II intermediate, III skilled (non-manual), III skilled (manual), IV partly skilled, V unskilled; those who reported as being in the armed forces with insufficient information to assign to these categories were recoded as missing. For the analyses, categories IV and V were combined due to small cell counts.

2) Family Adversity Index

A Family Adversity Index (FAI) was developed by the ALSPAC team using data from three developmental periods: pregnancy, 0-2 years, and 2-4 years of age. It is devised by summing items that reflected family-based risk factors. There are two versions of the FAI, with the long index comprising up to 18 items using as much information as possible at each time point, whereas the short index consists of 15 items for which data are potentially available across the three time periods. In this study, the long index from pregnancy, with individuals that have missing values in over 10 items excluded, was used.

These items include:

- Age of mother: at first pregnancy / child birth
- Housing:
  - Adequacy
  - Basic living
  - Defects/infestation
- No educational qualifications: achievement (mother or father)
- Financial status: financial difficulties
- Partner relationship
  - Status
  - Affection and aggression
  - Physical / emotional cruelty
  - No social support
- Family
  - Family size
  - Major problems (child in care / not with natural mother, or on at risk register)
- Social network
  - Emotional support
  - Practical / financial support
- Maternal psychopathology: affective (depression, anxiety, or suicide attempts)
- Substance abuse: drugs or alcohol (use of hard drugs, alcoholism, alcohol consumption)
- Crime:
  - In trouble with police
  - Convictions

3) Index of Multiple Deprivation

ALSPAC address data was initially collected upon recruitment of the mothers, and was historically updated in an ad-hoc manner based on participant self-reported residential moves. In child-based data, the family address is used unless the child informs ALSPAC of a new address for them. In this study we have used the English Index of Multiple Deprivation 2000 quintiles generated based on addresses recorded in the ALSPAC database at the age 10 clinic.

4) Depressive symptoms

Part of the depressive symptoms data were collected and managed using Research Electronic Data Capture (REDCap)^1^ hosted at the University of Bristol. REDCap is a secure, web-based software platform designed to support data capture for research studies. Further details on the administration of the questionnaires at research clinics or via post/email have been previously reported.^2^

5) Cardiometabolic phenotypes

At the age 24 clinic, standing height was measured using a Harpenden wall-mounted stadiometer and weight using a Tanita TBF-401A electronic body composition scales or electronic bathroom scales if the individual had a pacemaker. Body mass index (BMI; kg/m^2^) was computed using the formula: weight (kg) / height^2^ (m^2^). Prior to the calculation of BMI, outliers and implausible values of weight and height were identified by adapting the conditional growth percentiles method described in Yang and Hutcheon.^3^ First, using weight and height data from 10 time points between age 7 and 24, regression models with a quadratic age term was fitted separately for males and females, and data points that lie beyond ±4 standard deviations (SDs) from the individual’s conditional mean were flagged and manually reviewed. Flagged data points that likely reflect a data entry error either in weight or height were recoded as missing. Additionally, BMI values of <10 before the age 17 clinic or <15 at the age 17 or age 24 clinics or ≥60 were recoded as missing. BMI at 10 years was z-transformed before being included as a covariable in any of the models. Waist and hip circumferences were measured using Seca 201 body tension tape and repeated twice for accuracy. Waist-hip ratio was calculated as mean waist circumference (mm) / mean hip circumference (mm).

Seated blood pressure readings were taken using an Omron M6 upper arm blood pressure/pulse monitor after sitting and resting for two minutes. All blood pressure readings were taken using the individual’s right arm where possible. The derived average seated systolic and diastolic blood pressure variables that were calculated as the average of the first, second and (if collected) third seated blood pressure readings were used. Metabolic syndrome was defined based on the 2009 consensus definition from the International Diabetes Federation and the American Heart Association/National Heart, Lung, and Blood Institute.^4^ The presence of any three of five of the following risk factors constitutes a diagnosis of metabolic syndrome:

- Elevated waist circumference (≥94cm for white men, ≥90cm for non-white men, ≥80cm for women)
- Elevated triglycerides (≥1.7mmol/L)
- Reduced HDL-cholesterol (< 1.0mmol/L)
- Elevated blood pressure (systolic ≥130mmHg or diastolic ≥85mmHg)
- Elevated fasting glucose (≥5.6mmol/L)

Carotid intima-media thickness (cIMT) was assessed using an ultrasound machine (CardioHealth Panasonic and a 13.5MHz linear array broadband transducer; probe centre frequency 9.0MHz). The scanner automatically saves the scans and provides automated measurements and stops scanning once a predefined quality threshold is achieved. The mean of the mean right cIMT and mean left cIMT values was used. Carotid-femoral pulse wave velocity (cfPWV) was measured after resting five minutes in a semi-prone position using a Vicorder instrument (Skidmore Medical) with two blood pressure measurement channels and two Velcro pressure sensor cuffs (neck and leg). The measurement was repeated until three readings that were within 0.5m/sec of each other had been recorded. Waveforms quality was checked in order to ensure noise free and appropriate quality tracings. All cIMT and cfPWV measurements were done by sonographers who were trained, accredited and completed reproducibility measurements, and raw data were reviewed for outlying or abnormal values. Pregnant individuals were excluded.

6) Smoking and alcohol use

Smoking behaviour and alcohol use were assessed using questionnaires at the age 24 clinic. A composite smoking variable was derived by coding those who self-reported having smoked in the past 30 days and ≥1 cigarettes per day or ≥7 per week as smokers, and risky alcohol use was defined as an Alcohol Use Disorders Identification Test for Consumption (AUDIT-C) score ≥5.

7) ICD-10 depressive episode and generalised anxiety disorder diagnoses

ICD-10 depressive episode and ICD-10 generalised anxiety disorder diagnoses were derived from the self-administered Computerized Interview Schedule – Revised (CIS-R).^6^ The interview includes general questions to establish an overall picture of health, followed by 14 sections each scoring for a particular type of neurotic symptom, namely somatic symptoms, fatigue, concentration and forgetfulness, sleep problems, irritability, worry about physical health, depression, depressive ideas, worry, anxiety, phobias, panic, compulsions, and obsessions. An algorithm is then applied to derive a number of ICD-10 diagnoses for neuropsychiatric disorders.

8) Antidepressant medication use

Data on use of antidepressant medication for depression and anxiety, including names and types of antidepressants used, timeframes of usage, adherence, and reasons for stopping treatment were collected in the ‘Life @ 28’ Mental Health Treatments questionnaire section.^7^ A composite antidepressant prescription variable was derived based on self-reported prescription of a selective serotonin reuptake inhibitor (SSRI), a serotonin and norepinephrine reuptake inhibitor (SNRI), a tricyclic antidepressant (TCA), an atypical antidepressant or other antidepressant in the five year prior to the record. The anxiolytic prescription variable was based on self-reported prescription of an anxiolytic for depression and anxiety.

Biomarkers

1) Inflammatory proteins

Blood was collected in lithium heparin tubes and placed on ice until processing. Samples were spun at 1300g for 10 minutes at 4-5°C, then plasma was aliquoted from the tube and stored at -80°C. Target time from sample collection until plasma frozen was 90 minutes. Heparin-stored plasma samples collected from mothers in midlife (fasting) and offspring at approximately ages 9 (non-fasting) and 24 (fasting) were analysed using the Olink Target 96 Inflammation panel (Olink Analysis Service, Uppsala, Sweden). The Olink Target 96 Inflammation panel measures a selection of 92 proteins involved in inflammatory and immune response processes. Data were returned in Normalised Protein eXpression (NPX) values on a log­_2_ scale, which are calculated from Ct values with normalisation to minimise intra- and inter-assay variation. Further details on the proteins assayed and quality control procedures applied have been previously reported.^8^

Samples that did not pass Olink’s quality control were also excluded from the analysis. Proteins with ≥50% values below the limit of detection (LOD) were excluded (based on values reported in the data note; namely ARTN, Beta-NGF, FGF-23, FGF-5, GDNF, IL-1 alpha, IL-10RA, IL-13, IL-15RA, IL-17A, IL-2, IL-20, IL-20RA, IL-22 RA1, IL-24, IL-2RB, IL-33, IL-4, IL-5, LIF, NRT, NT-3, SIRT2, SLAMF1, TSLP), leaving 67 proteins for analysis (**Table S2**). NPX values beyond 5 standard deviations (SD) from the mean were recoded as missing. We checked for proteins or samples with >50% missingness, but all proteins and samples had sufficient data to be included. No further normalisation procedures were applied to the proteomic data prior to analysis.

2) NMR metabolomics

Metabolomic profiling was performed using a high-throughput ^1^H-NMR spectroscopy-based platform (Nightingale Health, Helsinki, Finland) using a standardised protocol and parameters described elsewhere.^9-11^ Briefly, 70 μL plasma and 70 μL sodium phosphate buffer (75 mM Na2HPO4, 0.08% sodium 3-(trimethylsilyl)propionate-2,2,3,3-d4, 0.04% sodium azide in 80%/20% H2O/D2O, pH 7.4) were mixed and transferred to 3 mm NMR tubes using an 8-channel Varispan Janus liquid handling robot (PerkinElmer). NMR spectra were acquired using Bruker Avance III HD 600MHz spectrometer equipped with a nitrogen-cooled triple resonance probe (CryoProbe Prodigy TCI) equipped with SampleJet auto-sampler with cooled (6°C) sample storage. Spectra were acquired using standardised parameters using two NMR experiments or ‘molecular windows’ to characterise lipoproteins, low molecular weight metabolites and lipids. Over 220 measures, including lipoproteins, low molecular weight metabolites and lipids were quantified per sample of EDTA plasma. For the present analysis, a subset of 71 metabolites capturing the majority of variation were selected to minimise redundancy of information^12^ (**Table S3**). Values beyond 5 SD from the mean were recoded as missing. We checked for metabolites or samples with >10% missingness, but all had sufficient data to be included. All metabolites were then log_2_-transformed.

3) Blood count and biochemistry markers

Full blood count was performed on fasting EDTA plasma samples at the NHS blood laboratory at Southmead Hospital, Bristol, UK. All biochemistry markers were also assayed on fasting EDTA plasma samples, with the exception of procollagen-3 N-terminal peptide (P3NP) and hyaluronic acid (HA), which were assayed on serum samples.

Insulin levels were measured using a commercially available electrochemiluminescence immunoassay (ECLIA) kit. Cholesterol, triglycerides, and HDL-cholesterol (HDL-c) were measured using enzymatic colorimetric tests, while glucose and C-reactive protein (CRP) were measured by the hexokinase method and particle enhanced immunoturbidimetric assay, respectively. LDL-cholesterol (LDL-c) was calculated using the Friedwald equation (LDL-c = total cholesterol - (HDL-c + triglycerides / 2.19)) and VLDL-cholesterol was calculated from triglyceride levels as VLDL-c = triglycerides / 2.19. GGT, ALT and AST were measured using commercially available enzymatic colorimetric assay kits.

The Homeostatic Model Assessment for Insulin Resistance (HOMA-IR) was calculated using the formula: HOMA1-IR = (fasting glucose (mmol/L) * fasting insulin (µU/mL)) / 22.5. All blood count and biochemistry marker values were ln(x+0.01)-transformed to reduce skewness. Details of all kits used can be found in **Table S4**.

*Statistical analysis*

Modelling of depressive symptom trajectories from childhood to early adulthood

A total of nine models were estimated to identify the most appropriate model specification – two one-class models to obtain starting values, five GMM with random intercepts and class-specific proportional random-effect variance-covariance matrix, one GMM with random intercepts and common random-effect variance-covariance matrix, and one GBTM. A total of 44 models were fitted to identify the most appropriate trajectory shape, then a one-class model with the best-fitting fractional polynomial was run to obtain starting values for estimation of the final model using grid search. Then classes in the final model were reordered using the permut function for ease of interpretation.

**Methods S2. R scripts for the latent class trajectory modelling and biomarker analyses**

\## LATENT CLASS TRAJECTORY MODELLING

set.seed(22780)

library(tidyverse)

library(lcmm)

library(LCTMtools)

library(gtools)

\# Run 1-class models

m1 <- hlme(smfq ~ ageyr + I(ageyr^2),

subject = 'id',

data = dep_long_inc,

maxiter = 500,

nproc = 10)

m1i <- hlme(smfq ~ ageyr + I(ageyr^2),

random = ~ 1,

subject = 'id',

data = dep_long_inc,

maxiter = 500,

nproc = 10)

m1s <- hlme(smfq ~ ageyr + I(ageyr^2),

random = ~ ageyr + I(ageyr^2),

subject = 'id',

data = dep_long_inc,

maxiter = 500,

nproc = 10)

\# Class enumeration

gmmis2 <- gridsearch(rep = 50, maxiter = 10, minit = m1i, cl = 10,

hlme(smfq ~ ageyr + I(ageyr^2),

random = ~ 1,

mixture = ~ ageyr + I(ageyr^2),

ng = 2, subject = 'id', nwg = TRUE,

data = dep_long_inc, maxiter = 500, nproc = 10,

verbose = TRUE))

gmmis3 <- gridsearch(rep = 50, maxiter = 10, minit = m1i, cl = 10,

hlme(smfq ~ ageyr + I(ageyr^2),

random = ~ 1,

mixture = ~ ageyr + I(ageyr^2),

ng = 3, subject = 'id', nwg = TRUE,

data = dep_long_inc, maxiter = 500, nproc = 10,

verbose = TRUE))

gmmis4 <- gridsearch(rep = 50, maxiter = 10, minit = m1i, cl = 10,

hlme(smfq ~ ageyr + I(ageyr^2),

random = ~ 1,

mixture = ~ ageyr + I(ageyr^2),

ng = 4, subject = 'id', nwg = TRUE,

data = dep_long_inc, maxiter = 500, nproc = 10,

verbose = TRUE))

gmmis5 <- gridsearch(rep = 50, maxiter = 10, minit = m1i, cl = 10,

hlme(smfq ~ ageyr + I(ageyr^2),

random = ~ 1,

mixture = ~ ageyr + I(ageyr^2),

ng = 5, subject = 'id', nwg = TRUE,

data = dep_long_inc, maxiter = 500, nproc = 10,

verbose = TRUE))

gmmis6 <- gridsearch(rep = 50, maxiter = 10, minit = m1i, cl = 10,

hlme(smfq ~ ageyr + I(ageyr^2),

random = ~ 1,

mixture = ~ ageyr + I(ageyr^2),

ng = 6, subject = 'id', nwg = TRUE,

data = dep_long_inc, maxiter = 500, nproc = 10,

verbose = TRUE))

\# Compare model fit and assess model adequacy of the best-fitting model

summarytable(m1i, gmmis2, gmmis3, gmmis4, gmmis5, gmmis6,

which = c("conv", "G", "npm", "loglik",

"BIC", "SABIC", "ICL",

"%class", "entropy"))

summaryplot(gmmis2, gmmis3, gmmis4, gmmis5)

LCTMtoolkit(gmmis4)

\# Test alternative model specification

gmmi4 <- gridsearch(rep = 50, maxiter = 10, minit = m1i, cl = 10,

hlme(smfq ~ ageyr + I(ageyr^2),

random = ~ 1,

mixture = ~ ageyr + I(ageyr^2),

ng = 4, subject = 'id', nwg = FALSE,

data = dep_long_inc, maxiter = 500, nproc = 10,

verbose = TRUE))

gbtm4 <- gridsearch(rep = 50, maxiter = 10, minit = m1, cl = 10,

hlme(smfq ~ ageyr + I(ageyr^2),

mixture = ~ ageyr + I(ageyr^2),

ng = 4, subject = 'id', nwg = FALSE,

data = dep_long_inc, maxiter = 500, nproc = 10,

verbose = TRUE))

summarytable(gbtm4, gmmi4, gmmis4,

which = c("conv", "G", "npm", "loglik",

"BIC", "SABIC", "ICL",

"%class", "entropy"))

LCTMtoolkit(gbtm4)

LCTMtoolkit(gmmi4)

LCTMtoolkit(gmmis4)

\# Refine polynomial

fp <- c("I(ageyr^-2)", "I(ageyr^-1)", "I(ageyr^-0.5)", "I(ageyr^0.5)",

"log(ageyr)", "ageyr", "I(ageyr^2)", "I(ageyr^3)")

fpcomb <- as.data.frame(combinations(8, 2, fp, set = FALSE, repeats.allowed = TRUE))

fpcomb <- fpcomb %>%

mutate(V2 = ifelse(V2 == V1, paste(V2, ":log(ageyr)", sep = ""), V2))

fp2 <- paste(fpcomb$V1, fpcomb$V2, sep = " + ")

m1fp1 <- map(fp, ~ {eval(parse(text=(paste0(

"hlme(smfq ~ ", .x,

", subject = 'id', data = dep_long_inc,

nproc = 10, maxiter = 500, verbose = TRUE)"))))})

mfp1B <- vector()

for (i in 1:8) {

mfp1B[[length(mfp1B) + 1]] <- paste("m1fp1[[", i, "]]", sep = "")

}

mfp1 <- map2(fp, mfp1B, ~ {eval(parse(text=(paste0(

"hlme(smfq ~ ", .x,

", mixture = ~ ", .x,

", ng = 4, subject = 'id', nwg = FALSE, B = ", .y,

", data = dep_long_inc,

nproc = 10, maxiter = 500, verbose = TRUE)"))))})

m1fp2 <- map(fp2, ~ {eval(parse(text=(paste0(

"hlme(smfq ~ ", .x,

", subject = 'id', data = dep_long_inc,

nproc = 10, maxiter = 500, verbose = TRUE)"))))})

mfp2B <- vector()

for (i in 1:36) {

mfp2B[[length(mfp2B) + 1]] <- paste("m1fp2[[", i, "]]", sep = "")

}

\# Note: some models failed to converge

\# Manually remove them from list of models for starting values before running the estimation below

mfp2 <- map2(fp2, mfp2B, ~ {eval(parse(text=(paste0(

"hlme(smfq ~ ", .x,

", mixture = ~ ", .x,

", ng = 4, subject = 'id', nwg = FALSE, B = ", .y,

", data = dep_long_inc,

nproc = 10, maxiter = 500, verbose = TRUE)"))))})

mfp_res <- map_df(c(mfp1, mfp2),

~ {return(data.frame(summarytable(.x,

which = c("conv", "loglik",

"BIC", "SABIC",

"entropy", "ICL",

"%class"))))})

\# Estimate final solution using the best-fitting polynomial

m1f <- hlme(smfq ~ I(ageyr^0.5) + log(ageyr),

subject = 'id',

data = dep_long_inc,

maxiter = 500,

nproc = 10)

mfin <- gridsearch(rep = 50, maxiter = 10, minit = m1f, cl = 10,

hlme(smfq ~ I(ageyr^0.5) + log(ageyr),

mixture = ~ I(ageyr^0.5) + log(ageyr),

ng = 4, subject = 'id', nwg = FALSE,

data = dep_long_inc, maxiter = 500, nproc = 10,

verbose = TRUE))

LCTMtoolkit(mfin)

\## BIOMARKER ANALYSES

set.seed(22780)

library(tidyverse)

library(limma)

library(broom)

\# Inflammatory proteins

olink_dep <- omics_dep %>%

select(id, sex, ageyr_24, mat_ed, mat_soc, bmi_10, bmi_24, smoking_24, auditc_24,

il8_24:olink_qc_warning_24, class, cisr_dep_24) %>%

mutate(across(c(il8_24:csf1_24), ~ ifelse(. > mean(., na.rm = TRUE) +

5 * sd(., na.rm = TRUE), NA,

ifelse(. < mean(., na.rm = TRUE) -

5 * sd(., na.rm = TRUE),

NA, .)))) %>%

mutate(varcount = rowSums(!is.na(.[, 10:101])),

c_ageyr_24 = as.numeric(scale(ageyr_24, center = TRUE, scale = FALSE)),

z_bmi_10 = as.numeric(scale(bmi_10, center = TRUE, scale = TRUE)),

z_bmi_24 = as.numeric(scale(bmi_24, center = TRUE, scale = TRUE)),

z_auditc_24 = as.numeric(scale(auditc_24, center = TRUE, scale = TRUE))) %>%

filter(!is.na(olink_qc_warning_24) & varcount >= 92/2 & !is.na(class) &

!is.na(sex) & !is.na(mat_ed) & !is.na(mat_soc) & !is.na(z_bmi_10)) %>%

mutate(across(c(sex, smoking_24, mat_ed, mat_soc, class, cisr_dep_24), as.factor)) %>%

select(-c(batch_24, plate_24, varcount))

\# Exclude proteins with >=50% below LOD

olink_dep <- olink_dep %>%

select(-c(artn_24, betangf_24, fgf23_24, fgf5_24, gdnf_24, il1alpha_24,

il10ra_24, il13_24, il15ra_24, il17a_24, il2_24, il20_24,

il20ra_24, il22ra1_24, il24_24, il2rb_24, il33_24, il4_24,

il5_24, lif_24, nrtn_24, nt3_24, sirt2_24, slamf1_24, tslp_24))

\# Exclude samples that did not pass Olink QC

olink_dep <- olink_dep %>%

filter(olink_qc_warning_24 == 1) %>%

select(-olink_qc_warning_24)

\# Basic model

npx_df <- olink_dep %>%

select(id, il8_24:csf1_24)

pheno_df <- olink_dep %>%

select(id, ageyr_24, c_ageyr_24, sex, mat_ed, mat_soc, bmi_10, z_bmi_10,

bmi_24, z_bmi_24, smoking_24, auditc_24, z_auditc_24, class,

cisr_dep_24)

rownames(npx_df) <- npx_df$id

npx_df <- npx_df %>% select(-id)

design <- model.matrix(~0 + class + sex, pheno_df)

fit <- lmFit(t(npx_df), design)

contr <- makeContrasts(class2 - class1, class3 - class1,

class4 - class1, levels = design)

fit2 <- contrasts.fit(fit, contr)

fit2 <- eBayes(fit2, proportion = 0.1)

top.table21 <- topTable(fit2, coef = 1, sort.by = "P", n = Inf,

adjust = "BH", confint = TRUE) %>%

rownames_to_column(var = "protein") %>%

mutate(FC = 2^(abs(logFC)))

top.table31 <- topTable(fit2, coef = 2, sort.by = "P", n = Inf,

adjust = "BH", confint = TRUE) %>%

rownames_to_column(var = "protein") %>%

mutate(FC = 2^(abs(logFC)))

top.table41 <- topTable(fit2, coef = 3, sort.by = "P", n = Inf,

adjust = "BH", confint = TRUE) %>%

rownames_to_column(var = "protein") %>%

mutate(FC = 2^(abs(logFC)))

\# Adjusted model

designa <- model.matrix(~0 + class + sex + mat_ed + mat_soc + z_bmi_10, pheno_df)

fita <- lmFit(t(npx_df), designa)

contra <- makeContrasts(class2 - class1, class3 - class1,

class4 - class1, levels = designa)

fit2a <- contrasts.fit(fita, contra)

fit2a <- eBayes(fit2a, proportion = 0.1)

top.table21a <- topTable(fit2a, coef = 1, sort.by = "P", n = Inf,

adjust = "BH", confint = TRUE) %>%

rownames_to_column(var = "protein") %>%

mutate(FC = 2^(abs(logFC)))

top.table31a <- topTable(fit2a, coef = 2, sort.by = "P", n = Inf,

adjust = "BH", confint = TRUE) %>%

rownames_to_column(var = "protein") %>%

mutate(FC = 2^(abs(logFC)))

top.table41a <- topTable(fit2a, coef = 3, sort.by = "P", n = Inf,

adjust = "BH", confint = TRUE) %>%

rownames_to_column(var = "protein") %>%

mutate(FC = 2^(abs(logFC)))

\# NMR metabolomics

nmr_dep <- omics_dep %>%

select(id, sex, ageyr_24, mat_ed, mat_soc, bmi_10, bmi_24, smoking_24, auditc_24,

vldld_24:gp_24, class, class_robust, cisr_dep_24) %>%

mutate(across(c(vldld_24:gp_24), ~ ifelse(. > mean(., na.rm = TRUE) +

5 * sd(., na.rm = TRUE), NA,

ifelse(. < mean(., na.rm = TRUE) -

5 * sd(., na.rm = TRUE),

NA, .)))) %>%

mutate(varcount = rowSums(!is.na(.[, c(10:66)])),

c_ageyr_24 = as.numeric(scale(ageyr_24, center = TRUE, scale = FALSE)),

z_bmi_10 = as.numeric(scale(bmi_10, center = TRUE, scale = TRUE)),

z_bmi_24 = as.numeric(scale(bmi_24, center = TRUE, scale = TRUE)),

z_auditc_24 = as.numeric(scale(auditc_24, center = TRUE, scale = TRUE))) %>%

filter(varcount >= 57 * 0.9 & !is.na(class) & !is.na(sex) & !is.na(mat_ed) &

!is.na(mat_soc) & !is.na(z_bmi_10)) %>%

mutate(across(c(vldld_24:gp_24), ~ log2(.))) %>%

mutate(across(c(sex, smoking_24, mat_ed, mat_soc, class, cisr_dep_24), as.factor)) %>%

select(-varcount)

\# Basic model

nmr_df <- nmr_dep %>%

select(id, vldld_24:gp_24)

pheno_df <- nmr_dep %>%

select(id, ageyr_24, c_ageyr_24, sex, mat_ed, mat_soc, bmi_10, z_bmi_10,

bmi_24, z_bmi_24, smoking_24, auditc_24, z_auditc_24, class,

cisr_dep_24)

rownames(nmr_df) <- nmr_df$id

nmr_df <- nmr_df %>% select(-id)

design <- model.matrix(~0 + class + sex, pheno_df)

fit <- lmFit(t(nmr_df), design)

contr <- makeContrasts(class2 - class1, class3 - class1,

class4 - class1, levels = design)

fit2 <- contrasts.fit(fit, contr)

fit2 <- eBayes(fit2)

top.table21 <- topTable(fit2, coef = 1, sort.by = "P", n = Inf,

adjust = "BH", confint = TRUE) %>%

rownames_to_column(var = "metabolite") %>%

mutate(FC = 2^(abs(logFC)))

top.table31 <- topTable(fit2, coef = 2, sort.by = "P", n = Inf,

adjust = "BH", confint = TRUE) %>%

rownames_to_column(var = "metabolite") %>%

mutate(FC = 2^(abs(logFC)))

top.table41 <- topTable(fit2, coef = 3, sort.by = "P", n = Inf,

adjust = "BH", confint = TRUE) %>%

rownames_to_column(var = "metabolite") %>%

mutate(FC = 2^(abs(logFC)))

\# Adjusted model

designa <- model.matrix(~0 + class + sex + mat_ed + mat_soc + z_bmi_10, pheno_df)

fita <- lmFit(t(nmr_df), designa)

contra <- makeContrasts(class2 - class1, class3 - class1,

class4 - class1, levels = designa)

fit2a <- contrasts.fit(fita, contra)

fit2a <- eBayes(fit2a)

top.tablea <- topTable(fit2a, sort.by = "F", n = Inf, adjust = "BH") %>%

rownames_to_column(var = "metabolite") %>%

mutate(metabolite = sub("_24", "", metabolite))

top.table21a <- topTable(fit2a, coef = 1, sort.by = "P", n = Inf,

adjust = "BH", confint = TRUE) %>%

rownames_to_column(var = "metabolite") %>%

mutate(FC = 2^(abs(logFC)))

top.table31a <- topTable(fit2a, coef = 2, sort.by = "P", n = Inf,

adjust = "BH", confint = TRUE) %>%

rownames_to_column(var = "metabolite") %>%

mutate(FC = 2^(abs(logFC)))

top.table41a <- topTable(fit2a, coef = 3, sort.by = "P", n = Inf,

adjust = "BH", confint = TRUE) %>%

rownames_to_column(var = "metabolite") %>%

mutate(FC = 2^(abs(logFC)))

\# Blood count and biochemistry

blood_dep <- omics_dep %>%

select(id, sex, ageyr_24, auditc_24, smoking_24, mat_ed, mat_soc, bmi_10, bmi_24,

wbc_24:p3np_24, ha_24, class, cisr_dep_24) %>%

mutate(across(c(wbc_24:p3np_24, ha_24), ~ ifelse(. > mean(., na.rm = TRUE) + 5 *

sd(., na.rm = TRUE), NA,

ifelse(. < mean(., na.rm = TRUE) - 5 *

sd(., na.rm = TRUE), NA, .)))) %>%

mutate(homa_ir_24 = (insulin_24 * glucose_24) / 22.5,

ast_alt_24 = ast_24 / alt_24,

varcount = rowSums(!is.na(.[, c(10:35)])),

c_ageyr_24 = ageyr_24 - 24,

z_bmi_10 = as.numeric(scale(bmi_10, center = TRUE, scale = TRUE)),

z_bmi_24 = as.numeric(scale(bmi_24, center = TRUE, scale = TRUE)),

z_auditc_24 = as.numeric(scale(auditc_24, center = TRUE, scale = TRUE))) %>%

filter(varcount >= 1 & !is.na(class) & !is.na(sex) & !is.na(mat_ed) & !is.na(mat_soc) &

!is.na(z_bmi_10)) %>%

mutate(across(c(wbc_24:p3np_24, ha_24, homa_ir_24:ast_alt_24), ~

log(. + 0.01), .names = "ln_{col}")) %>%

mutate(across(c(sex, smoking_24, mat_ed, mat_soc, class, cisr_dep_24),

as.factor)) %>%

select(-varcount)

\# Basic model

m1 <- blood_dep %>%

gather(marker, value, c(ln_wbc_24:ln_ast_alt_24)) %>%

group_by(marker) %>%

do(tidy(lm(value ~ class + sex, data = .),

conf.int = TRUE, conf.level = 0.95)) %>%

filter(term == "class2" | term == "class3" | term == "class4") %>%

cbind(., adj.p = p.adjust(.$p.value, "BH")) %>%

mutate(marker = sub("_24", "", marker),

OR = exp(estimate) - 0.01,

OR.lci = exp(conf.low) - 0.01,

OR.uci = exp(conf.high) - 0.01,

model = "Basic",

label = case_when(adj.p < 0.1 ~ "*"))

m2 <- blood_dep %>%

gather(marker, value, c(ln_wbc_24:ln_ast_alt_24)) %>%

group_by(marker) %>%

do(tidy(lm(value ~ class + sex + mat_ed + mat_soc + z_bmi_10, data = .),

conf.int = TRUE, conf.level = 0.95)) %>%

filter(term == "class2" | term == "class3" | term == "class4") %>%

cbind(., adj.p = p.adjust(.$p.value, "BH")) %>%

mutate(marker = sub("_24", "", marker),

OR = exp(estimate) - 0.01,

OR.lci = exp(conf.low) - 0.01,

OR.uci = exp(conf.high) - 0.01,

model = "Adjusted",

label = case_when(adj.p < 0.1 ~ "*"))

**Methods S3. Sensitivity analyses**

In the primary analysis, one of the criteria used for model selection was an average posterior probability of ≥0.7 (which is commonly accepted in the literature), which applies a threshold to the level of certainty regarding latent class assignment. We decided to run the primary analysis using the categorical class membership without any further restrictions to maximise sample size and for ease of interpretation.

However, to address the issue of uncertainty potentially carrying over the subsequent analyses, we conducted two sets of sensitivity analyses:

1) The first set of analyses was conducted by limiting the sample to only individuals with their modal posterior probability ≥0.7 which further limits uncertainty, and
2) The second by using the estimated posterior probabilities instead of the assigned class membership in the models.

The results of these sensitivity analyses are presented below.

**Sensitivity analysis 1 – restricting the sample to those with modal posterior probability ≥0.7**

Applying this restriction drops 2571 participants, leaving n=7024.

The pattern of associations of class membership with sociodemographic variables are the same as that observed in the primary analysis:

|  | Low-stable  (n=5254) | Adolescent-limited  (n=639) | Adolescent-persistent  (n=519) | Adulthood-onset  (n=612) | *p* |
| --- | --- | --- | --- | --- | --- |
| Sex: Female | 2458 (46.8%) | 468 (73.2%) | 420 (80.9%) | 402 (65.7%) | <0.001 |
| Ethnicity: Non-white | 191 (4.1%) | 19 (3.3%) | 26 (5.7%) | 26 (4.9%) | 0.215 |
| Maternal education |  |  |  |  | <0.001 |
| CSE or none | 639 (13.6%) | 56 (9.5%) | 85 (18.2%) | 73 (13.4%) |  |
| Vocational | 427 (9.1%) | 36 (6.1%) | 39 (8.4%) | 37 (6.8%) |  |
| O-level | 1640 (35.0%) | 215 (36.6%) | 183 (39.3%) | 187 (34.3%) |  |
| A-level | 1212 (25.9%) | 162 (27.6%) | 107 (23.0%) | 154 (28.3%) |  |
| Degree | 769 (16.4%) | 118 (20.1%) | 52 (11.2%) | 94 (17.2%) |  |
| Maternal occupational social class | | | | | 0.488 |
| I – highest | 302 (7.4%) | 44 (8.9%) | 21 (5.3%) | 33 (7.2%) |  |
| II | 1378 (33.9%) | 176 (35.4%) | 131 (32.9%) | 162 (35.2%) |  |
| III (non-manual) | 1734 (42.7%) | 204 (41.0%) | 168 (42.2%) | 191 (41.5%) |  |
| III (manual) | 278 (6.8%) | 32 (6.4%) | 27 (6.8%) | 34 (7.4%) |  |
| IV or V^1^ – lowest | 369 (9.1%) | 41 (8.2%) | 51 (12.8%) | 40 (8.7%) |  |
| English IMD 2000 quintile | | | | | 0.406 |
| 1 – least deprived | 1366 (33.6%) | 214 (39.4%) | 119 (31.3%) | 164 (33.5%) |  |
| 2 | 827 (20.3%) | 96 (17.7%) | 85 (22.4%) | 102 (20.8%) |  |
| 3 | 747 (18.4%) | 95 (17.5%) | 65 (17.1%) | 91 (18.6%) |  |
| 4 | 599 (14.7%) | 77 (14.2%) | 61 (16.1%) | 79 (16.1%) |  |
| 5 – most deprived | 531 (13.0%) | 61 (11.2%) | 50 (13.2%) | 54 (11.0%) |  |
| Family Adversity Index | 1 [0, 1] | 1 [0, 2] | 1 [0, 2] | 1 [0, 2] | <0.001 |
| BMI at age 10 | 17.38 [15.96, 19.60] | 17.60 [16.10, 19.96] | 18.32 [16.36, 20.97] | 17.44 [15.98, 19.96] | <0.001 |
| Number of SMFQ measurements | 5 [3, 8] | 8 [6, 9] | 6 [4, 8] | 7 [5, 9] | <0.001 |

Associations with anthropometric, cardiometabolic and psychiatric outcomes:

|  |  | Adolescent-limited | | Adolescent-persistent | | Adulthood-onset |
| --- | --- | --- | --- | --- | --- | --- |
| Outcomes assessed at age 24 | n | Adjusted unstandardised regression coefficient (SE) | | | | |
| cIMT (continuous) | 1337 | 0.0017 (0.0037) | -0.0070 (0.0053) | | -0.0047 (0.0042) | |
| cfPWV (continuous) | 1467 | -0.0600 (0.0840) | -0.1252 (0.1236) | | 0.0538 (0.0937) | |
|  |  | Adjusted odds ratio (95% CI) | | | | |
| Smoking | 2313 | **1.70 (1.30-2.23)** | **2.73 (1.92-3.86)** | | **1.85 (1.39-2.45)** | |
| AUDIT-C score ≥ 5 | 2294 | 0.95 (0.74-1.23) | **0.58 (0.41-0.81)** | | 0.76 (0.58-0.99) | |
| BMI ≥ 30kg/m^2^ | 2313 | 1.45 (0.95-2.19) | **2.15 (1.29-3.51)** | | 1.47 (0.93-2.28) | |
| Elevated waist circumference | 2292 | 1.34 (0.99-1.80) | 1.31 (0.87-1.95) | | 1.14 (0.81-1.59) | |
| Triglycerides ≥1.7mmol/L | 1906 | 1.56 (0.92-2.56) | 1.29 (0.56-2.66) | | 1.43 (0.82-2.39) | |
| HDL <1.0mmol/L | 1906 | 0.92 (0.62-1.34) | 1.31 (0.79-2.09) | | 1.17 (0.79-1.71) | |
| SBP ≥ 130mmHg | 2330 | 0.89 (0.56-1.37) | 0.74 (0.34-1.45) | | 0.85 (0.53-1.31) | |
| DBP ≥ 85mmHg | 2330 | 1.31 (0.51-2.95) | 0.85 (0.19-2.60) | | 1.51 (0.59-3.37) | |
| Fasting glucose ≥5.6mmol/L | 1906 | 0.77 (0.54-1.08) | 1.04 (0.63-1.67) | | 0.97 (0.69-1.35) | |
| Metabolic syndrome | 2336 | 1.10 (0.62-1.86) | 0.84 (0.36-1.76) | | 0.83 (0.43-1.50) | |
| ICD-10 depressive episode | 2310 | **4.76 (2.93-7.71)** | **30.75 (19.44-49.38)** | | **12.00 (7.83-18.61)** | |
| ICD-10 GAD | 2303 | **4.25 (2.64-6.78)** | **22.48 (14.25-35.83)** | | **7.77 (5.04-12.02)** | |
| Outcomes assessed at age 28 |  |  |  | |  | |
| Prescribed antidepressants in past 5y | 2382 | **3.34 (2.41-4.61)** | **8.36 (5.87-11.92)** | | **5.51 (4.01-7.56)** | |
| Prescribed anxiolytics in past 5y | 2414 | **2.74 (1.37-5.25)** | **7.79 (4.18-14.36)** | | **5.19 (2.84-9.38)** | |

Notes: Regression models were adjusted for sex, maternal education, maternal occupational social class and BMI at age 10. Effect estimates presented are unstandardised regression coefficients for continuous outcomes and odds ratios (95% confidence intervals) for binary outcomes. The reference group for all analyses is the low-stable trajectory. Text in bold indicates evidence for the association after FDR correction of p-values.

Metabolic syndrome was defined based on the 2009 consensus definition from the International Diabetes Federation and the American Heart Association/National Heart, Lung, and Blood Institute, i.e. the presence of any three of the following five risk factors: elevated waist circumference (≥94cm for white men, ≥90cm for non-white men, ≥80cm for women); elevated triglycerides (≥1.7mmol/L), reduced high-density lipoprotein-cholesterol (<1.0mmol/L), elevated blood pressure (systolic ≥130mmHg or diastolic ≥85mmHg), and elevated fasting glucose (≥5.6mmol/L).

Abbreviations: AUDIT-C – Alcohol Use Disorders Identification Test for Consumption; BMI – body mass index; cfPWV – carotid-femoral pulse wave velocity; cIMT – carotid intima-media thickness; DBP – diastolic blood pressure; GAD – generalised anxiety disorder; HDL – high-density lipoprotein; ICD-10 – International Classification of Diseases 10^th^ Revision; SBP – systolic blood pressure

Inflammation proteins analysis (n=1748):

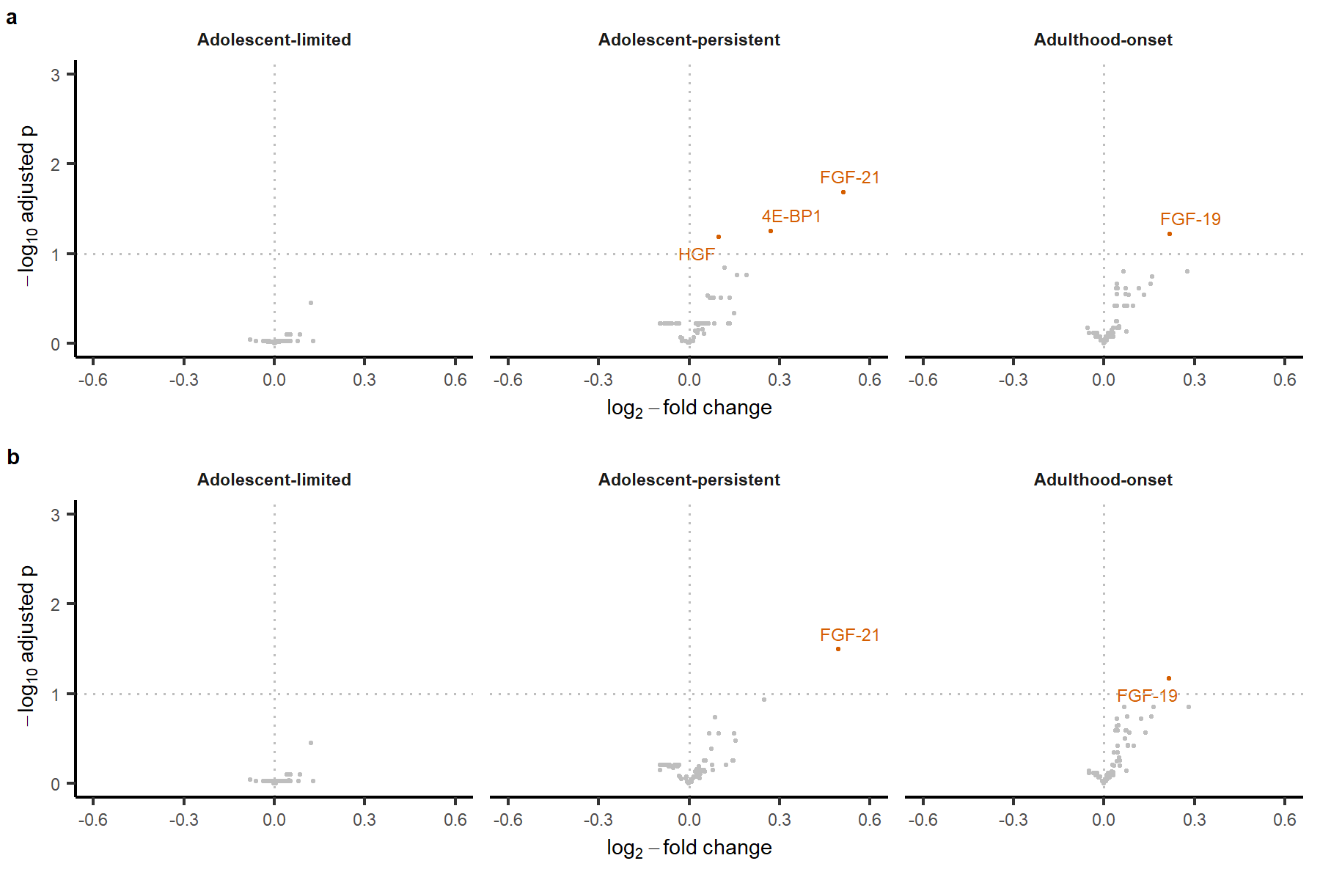

NMR metabolomics analysis (n=1906):

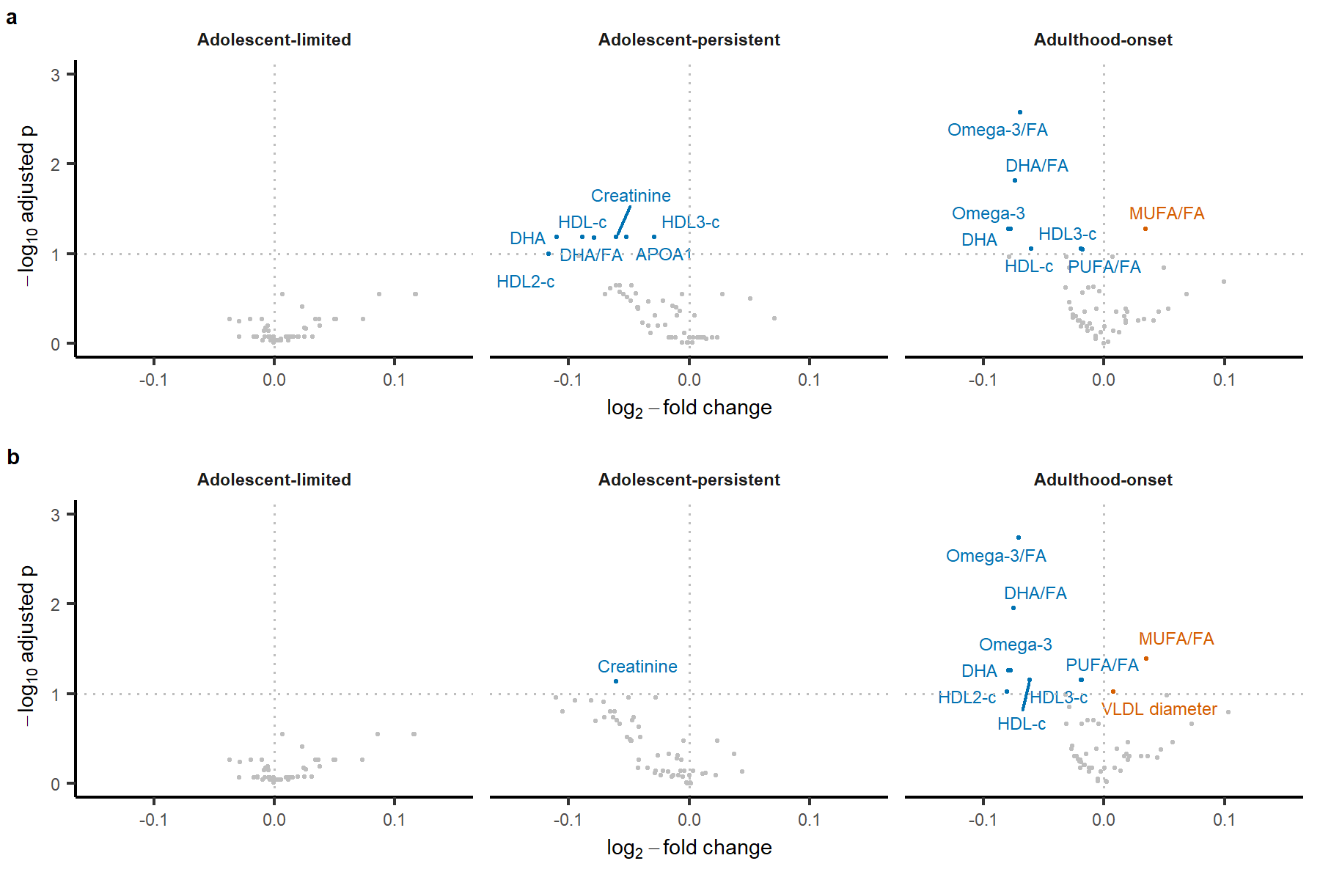

Full blood count and clinical biochemistry markers (n=1911):

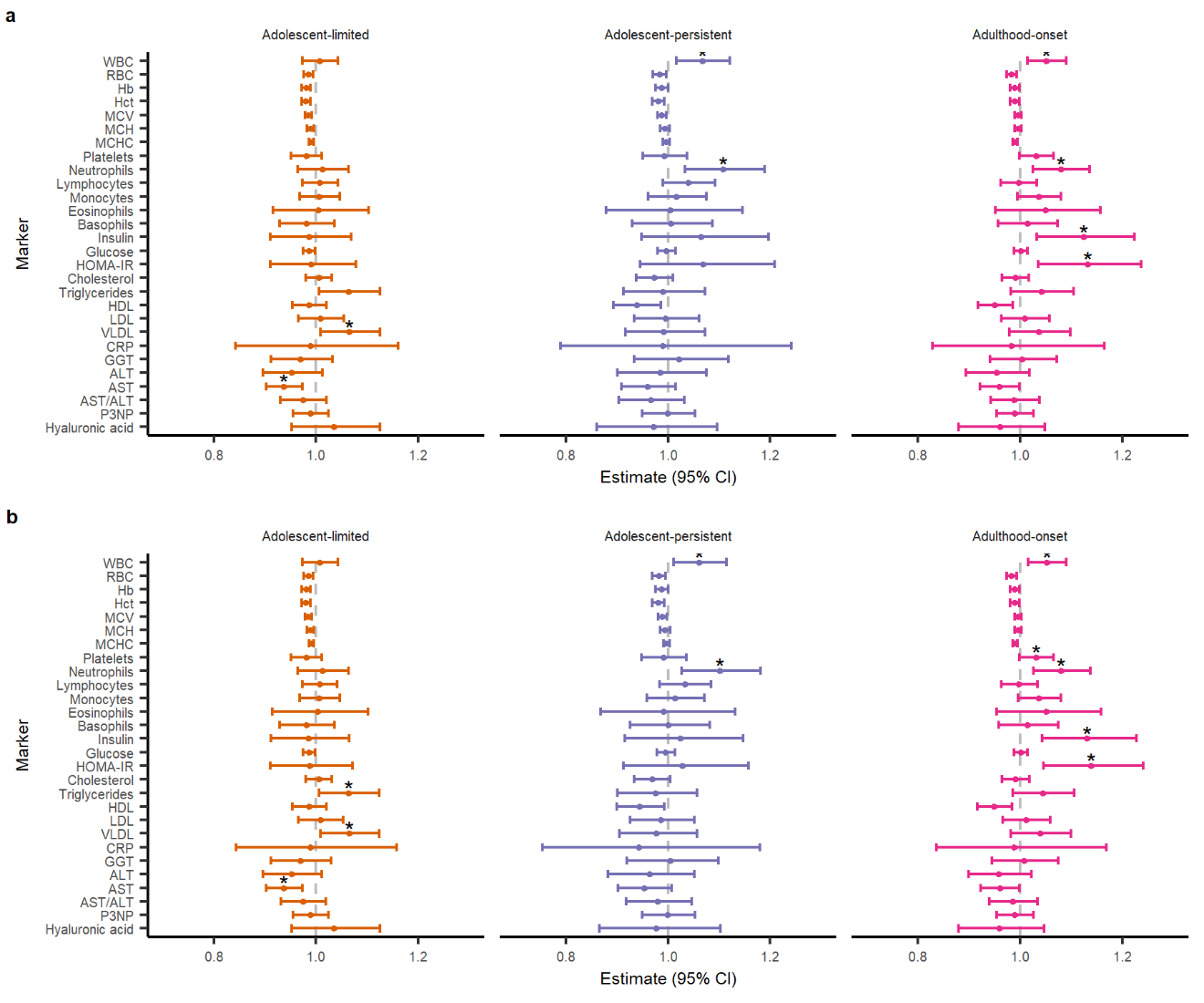

**Sensitivity analysis 2 – using individual posterior probabilities in the models**

Since the posterior probabilities for all four trajectories are collinear, the posterior probabilities of the adolescent-limited, adolescent-persistent and adulthood-onset trajectories as separate predictors in each of the models.

Note that the interpretation of the results from this set of sensitivity analyses is different to the ones carried out with trajectory classes as a single categorical variable, i.e. rather than comparing the depression-related trajectories to the low-stable trajectory, the coefficients represent comparisons between those assigned to a particular trajectory and everyone else in the sample, and therefore the effect sizes are not directly comparable to the those reported in the other analyses or among themselves.

Associations with anthropometric, cardiometabolic and psychiatric outcomes:

|  |  | Adolescent-limited | | Adolescent-persistent | | Adulthood-onset |
| --- | --- | --- | --- | --- | --- | --- |
| Outcomes assessed at age 24 | N | Adjusted unstandardised regression coefficient (SE) | | | | |
| cIMT (continuous) | 1709 | 0.0035 (0.0037) | -0.0057 (0.0050) | | -0.0034 (0.0040) | |
| cfPWV (continuous) | 1860 | -0.0433 (0.0796) | -0.1431 (0.1110) | | 0.0041 (0.0874) | |
|  |  | Adjusted odds ratio (95% CI) | | | | |
| Smoking | 2731 | **1.80 (1.36-2.36)** | **3.05 (2.15-4.31)** | | **2.06 (1.55-2.74)** | |
| AUDIT-C score ≥ 5 | 2704 | 0.99 (0.77-1.29) | **0.61 (0.43-0.85)** | | 0.81 (0.62-1.06) | |
| BMI ≥ 30kg/m^2^ | 2731 | 1.55 (1.01-2.35) | **2.22 (1.33-3.63)** | | 1.40 (0.87-2.19) | |
| Elevated waist circumference | 2707 | 1.42 (1.04-1.93) | 1.32 (0.88-1.98) | | 1.17 (0.83-1.63) | |
| Triglycerides ≥1.7mmol/L | 2241 | 1.50 (0.87-2.50) | 1.29 (0.58-2.63) | | 1.59 (0.92-2.67) | |
| HDL <1.0mmol/L | 2241 | 0.89 (0.60-1.30) | 1.20 (0.72-1.93) | | 1.28 (0.87-1.88) | |
| SBP ≥ 130mmHg | 2751 | 0.88 (0.55-1.36) | 0.63 (0.30-1.24) | | 0.97 (0.62-1.48) | |
| DBP ≥ 85mmHg | 2751 | 1.40 (0.54-3.29) | 0.76 (0.17-2.44) | | 1.55 (0.59-3.64) | |
| Fasting glucose ≥5.6mmol/L | 2241 | 0.84 (0.59-1.17) | 1.03 (0.63-1.63) | | 0.95 (0.67-1.34) | |
| Metabolic syndrome | 2759 | 1.34 (0.70-2.10) | 0.86 (0.38-1.77) | | 1.18 (0.65-2.04) | |
| ICD-10 depressive episode | 2729 | **5.58 (3.46-9.00)** | **36.58 (23.35-58.05)** | | **13.31 (8.63-20.75)** | |
| ICD-10 GAD | 2719 | **4.53 (2.79-7.33)** | **28.96 (18.46-45.87)** | | **8.68 (5.55-13.63)** | |
| Outcomes assessed at age 28 |  |  |  | |  | |
| Prescribed antidepressants in past 5y | 2792 | **3.41 (2.44-4.77)** | **9.20 (6.45-13.11)** | | **6.90 (4.99-9.55)** | |
| Prescribed anxiolytics in past 5y | 2828 | **2.59 (1.23-5.25)** | **8.81 (4.69-16.40)** | | **5.88 (3.11-11.04)** | |

Notes: Regression models were adjusted for sex, maternal education, maternal occupational social class and BMI at age 10. Effect estimates presented are unstandardised regression coefficients for continuous outcomes and odds ratios (95% confidence intervals) for binary outcomes. The reference group for all analyses is the low-stable trajectory. Text in bold indicates evidence for the association after FDR correction of p-values.

Metabolic syndrome was defined based on the 2009 consensus definition from the International Diabetes Federation and the American Heart Association/National Heart, Lung, and Blood Institute, i.e. the presence of any three of the following five risk factors: elevated waist circumference (≥94cm for white men, ≥90cm for non-white men, ≥80cm for women); elevated triglycerides (≥1.7mmol/L), reduced high-density lipoprotein-cholesterol (<1.0mmol/L), elevated blood pressure (systolic ≥130mmHg or diastolic ≥85mmHg), and elevated fasting glucose (≥5.6mmol/L).

Abbreviations: AUDIT-C – Alcohol Use Disorders Identification Test for Consumption; BMI – body mass index; cfPWV – carotid-femoral pulse wave velocity; cIMT – carotid intima-media thickness; DBP – diastolic blood pressure; GAD – generalised anxiety disorder; HDL – high-density lipoprotein; ICD-10 – International Classification of Diseases 10^th^ Revision; SBP – systolic blood pressure

Inflammation proteomics (n=2059):

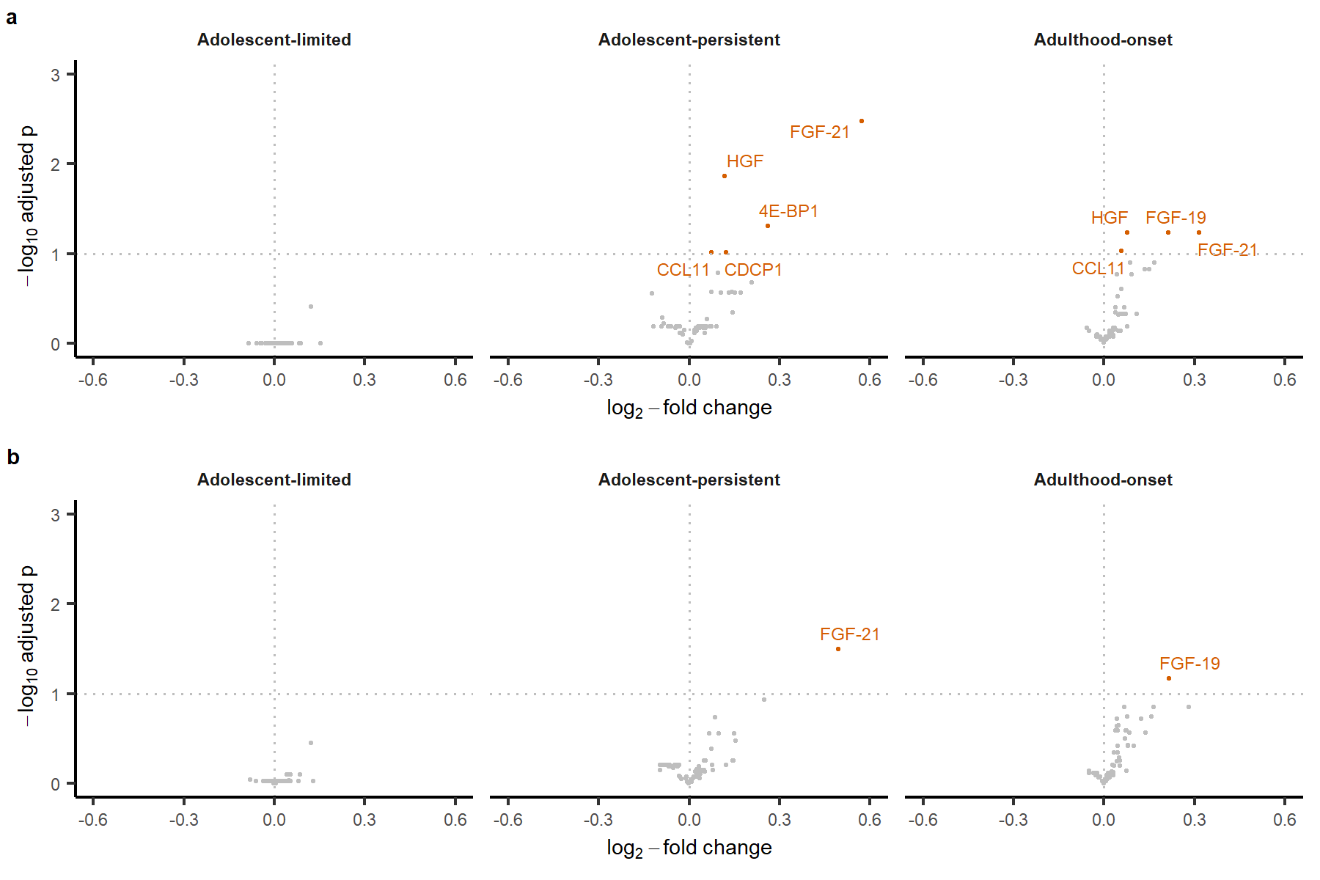

NMR metabolomics (n=2240):

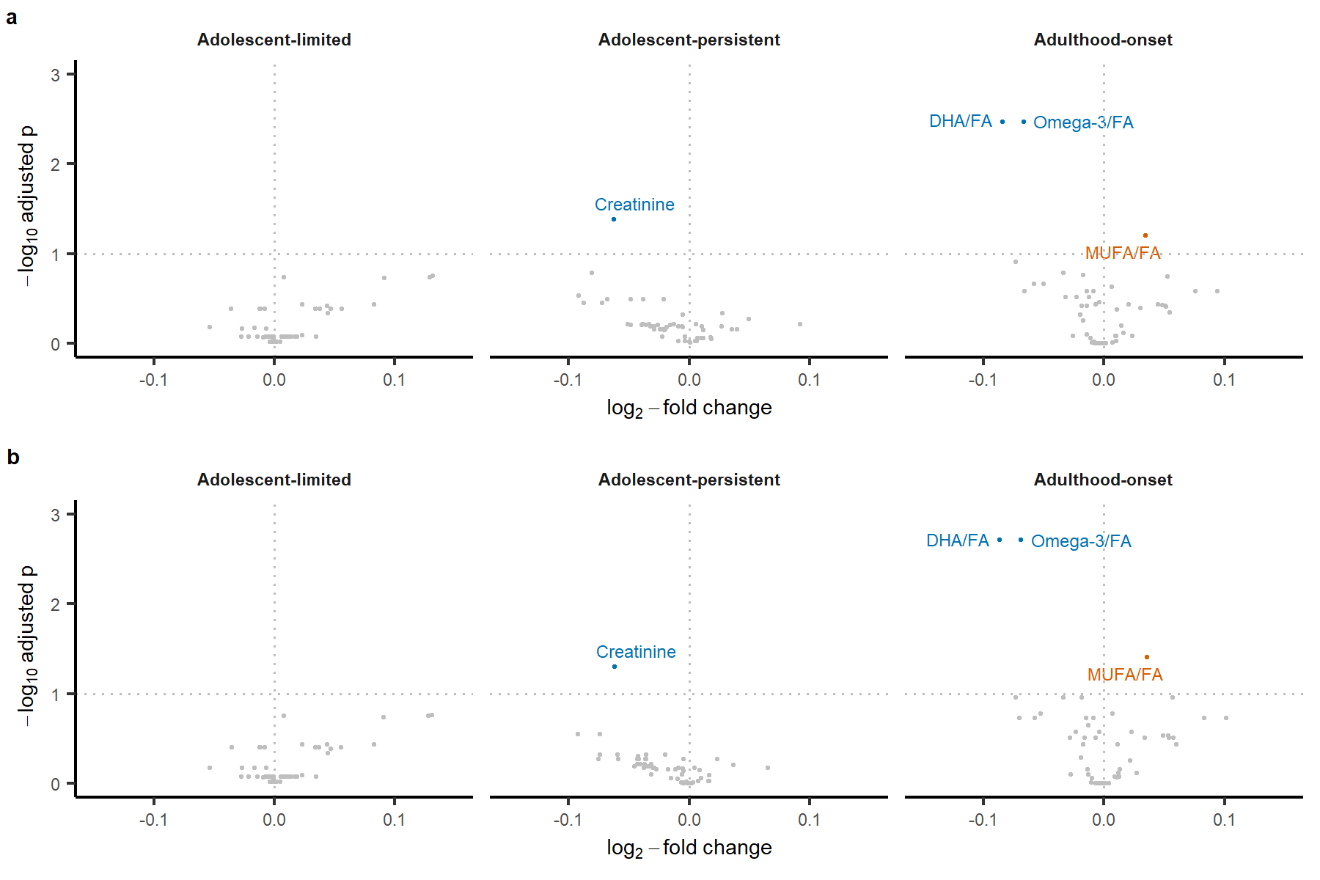

Full blood count and clinical biochemistry markers (n=2246):

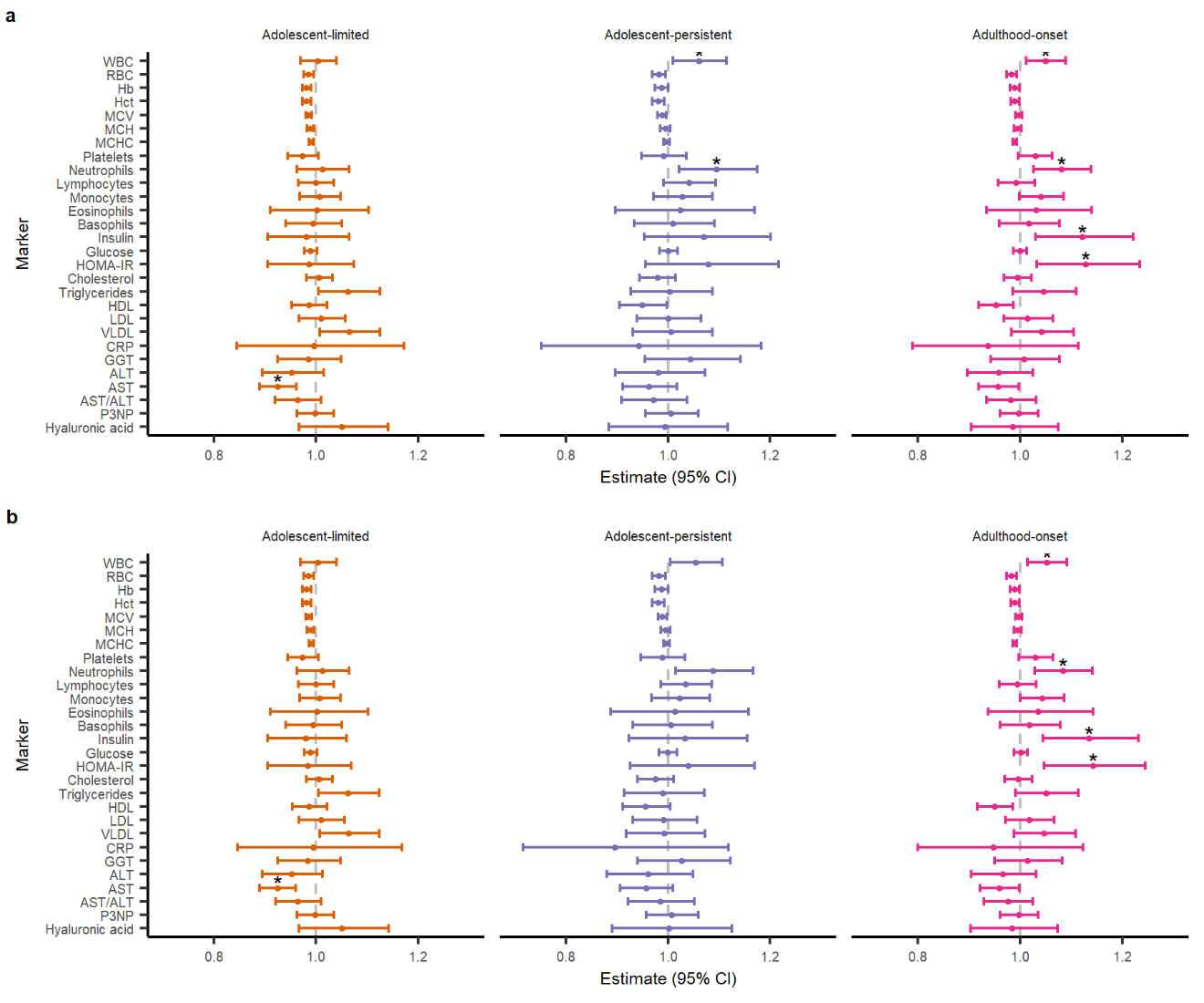

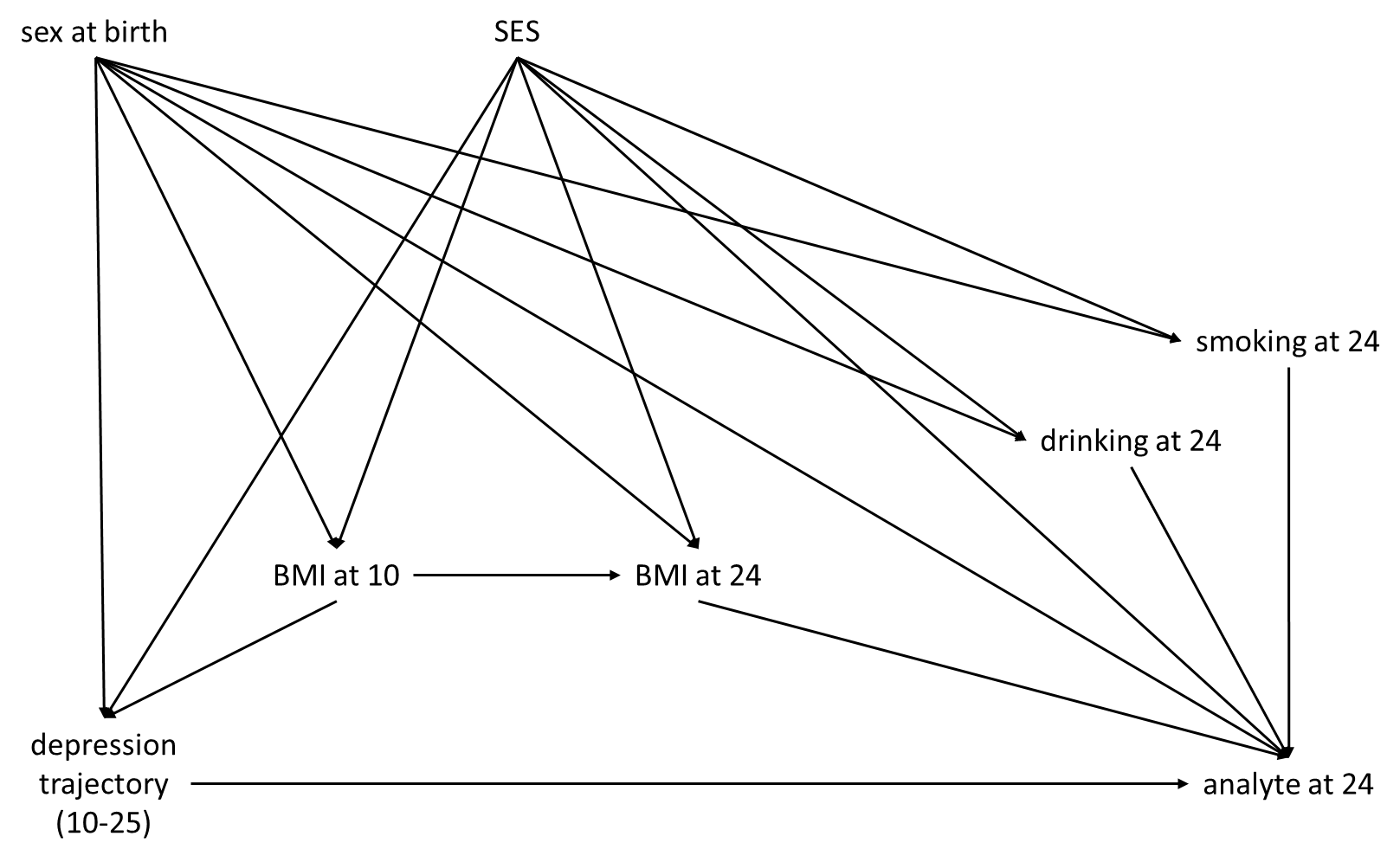

**Figure S1. Directed acyclic graph for confounder assessment.**

Final set of covariates included in the models: sex, SES (maternal education and maternal occupational social class), and BMI at 10.

Children alive at

1 year of age

(n=14901)

Posterior probabilities and class membership estimated

(n=9595)

Proteomics data available

(n=2970)

Metabolomics data available

(n=3203)

≥1 blood count or biochemistry measure

(n=3196)

Complete covariables

(n=2059)

Complete covariables

(n=2240)

Complete covariables

(n=2246)

≥3 measurements of SMFQ

(n=7302)

**Figure S2. Flow diagram illustrating sample selection for the latent class trajectory modelling and the biomarker analyses.**

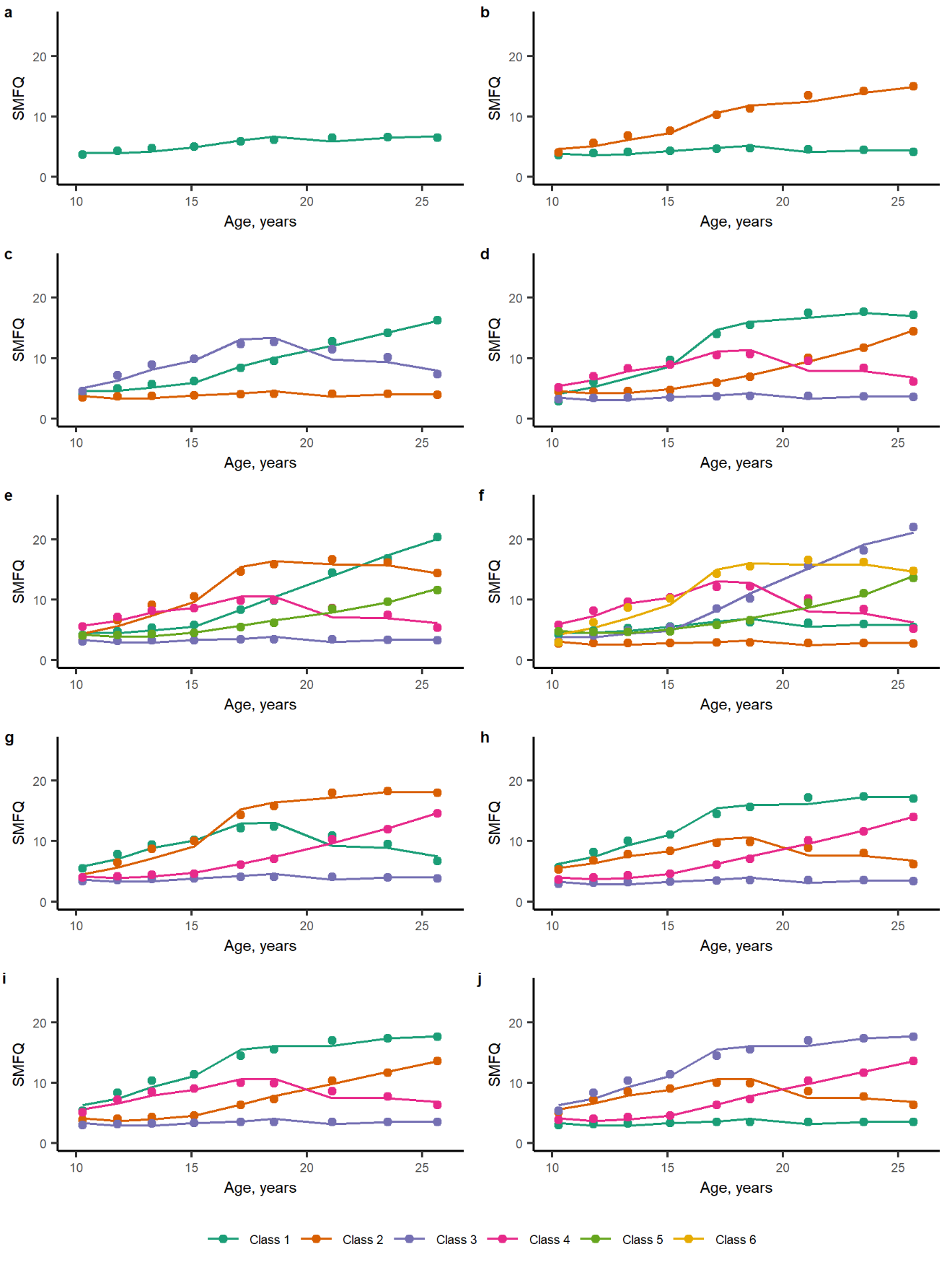

**Figure S3. Plots of weighted marginal predictions for models tested.**

Figures **a** is from the 1-class GMM, **b-f** from 2- to 6-class GMMs with class-specific variance-covariance matrices fitted at the class enumeration stage, **g** the 4-class GMM with common variance-covariance matrix, **h** the 4-class GBTM, **i** the 4-class GBTM with optimal trajectory shape, and **j** the 4-class GBTM with optimal trajectory shape with classes reordered. The points reflect predicted values and the lines reflect observed values.

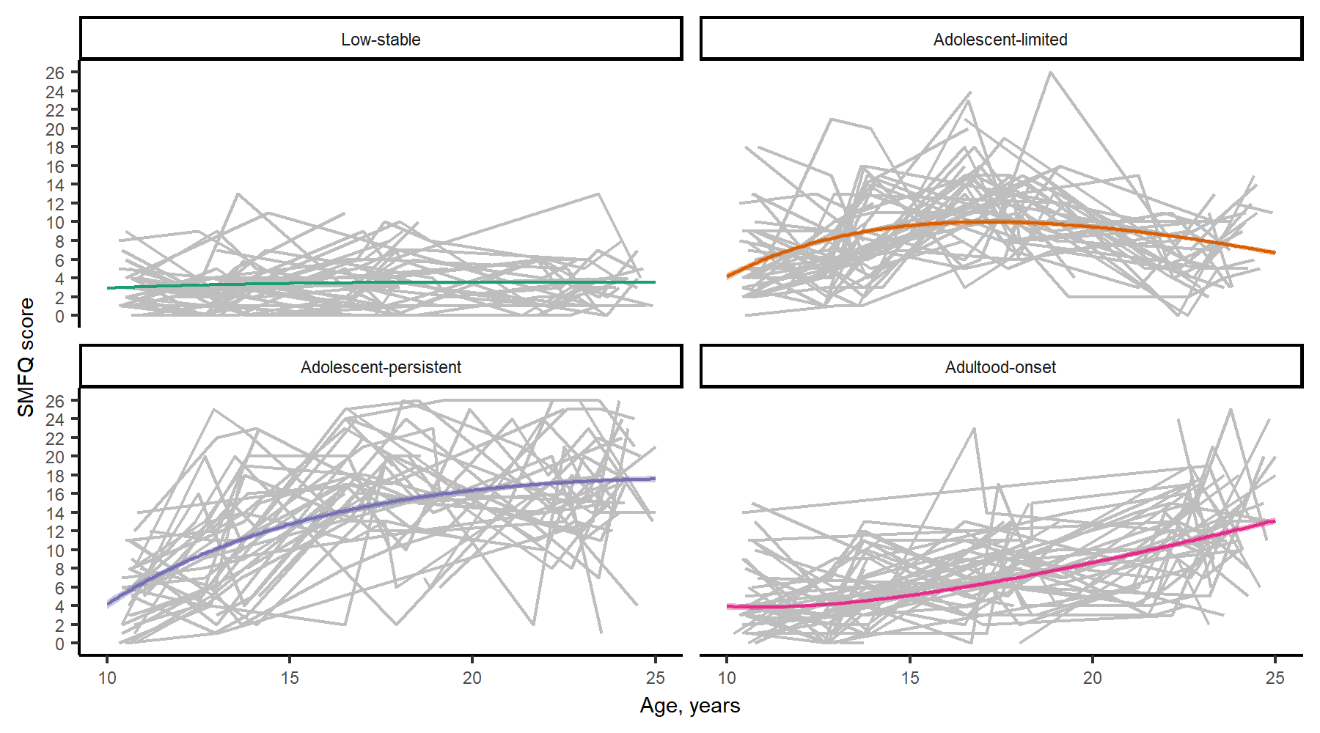

**Figure S4. Plots of predicted trajectories from the final model alongside individual trajectories.**

This plot shows the individual trajectories of a randomly sampled 50 individuals (in grey) against the predicted trajectories for each class (coloured).

**Table S1. Details of SMFQ time points included in the trajectory modelling.**

| Time point | Clinic/questionnaire | n | Age  (mean ± SD) | Total SMFQ score (mean ± SD) |
| --- | --- | --- | --- | --- |
| 1 | Clinic | 7387 | 10.6±0.27 | 4.05±3.52 |
| 2 | Clinic | 6743 | 12.8±0.23 | 3.98±3.87 |
| 3 | Clinic | 6049 | 13.8±0.21 | 4.92±4.49 |
| 4 | Questionnaire | 5072 | 16.7±0.24 | 5.89±5.64 |
| 5 | Clinic | 4487 | 17.8±0.46 | 6.59±5.25 |
| 6 | Questionnaire | 3341 | 18.7±0.49 | 6.81±5.91 |
| 7 | Questionnaire | 3396 | 22.0±0.52 | 5.67±5.57 |
| 8 | Questionnaire | 3970 | 22.9±0.53 | 6.19±5.53 |
| 9 | Questionnaire | 4082 | 23.9±0.52 | 7.01±6.06 |
| 10 | Questionnaire | 4129 | 25.8±0.51 | 6.82±6.40 |

**Table S2. List of 67 analysed inflammation proteomic markers.**

| Full name | Short name | UniprotID |
| --- | --- | --- |
| Eukaryotic translation initiation factor 4E-binding protein 1 | 4E-BP1 | Q13541 |
| Adenosine Deaminase | ADA | P00813 |
| Axin-1 | AXIN1 | O15169 |
| Caspase-8 | CASP-8 | Q14790 |
| Eotaxin | CCL11 | P51671 |
| C-C motif chemokine 19 | CCL19 | Q99731 |
| C-C motif chemokine 20 | CCL20 | P78556 |
| C-C motif chemokine 23 | CCL23 | P55773 |
| C-C motif chemokine 25 | CCL25 | O15444 |
| C-C motif chemokine 28 | CCL28 | Q9NRJ3 |
| C-C motif chemokine 3 | CCL3 | P10147 |
| C-C motif chemokine 4 | CCL4 | P13236 |
| Natural killer cell receptor 2B4 | CD244 | Q9BZW8 |
| CD40L receptor | CD40 | P25942 |
| T-cell surface glycoprotein CD5 | CD5 | P06127 |
| T cell surface glycoprotein CD6 isoform | CD6 | P30203 |
| T-cell surface glycoprotein CD8 alpha chanin | CD8A | P01732 |
| CUB domain-containing protein 1 | CDCP1 | Q9H5V8 |
| Macrophage colony-stimulating factor 1 | CSF-1 | P09603 |
| Cystatin D | CST5 | P28325 |
| Fractalkine | CX3CL1 | P78423 |
| C-X-C motif chemokine 1 | CXCL1 | P09341 |
| C-X-C motif chemokine 10 | CXCL10 | P02778 |
| C-X-C motif chemokine 11 | CXCL11 | O14625 |
| C-X-C motif chemokine 5 | CXCL5 | P42830 |
| C-X-C motif chemokine 6 | CXCL6 | P80162 |
| C-X-C motif chemokine 9 | CXCL9 | Q07325 |
| Delta and Notch-like epidermal growth factor-related receptor | DNER | Q8NFT8 |
| Protein S100-A12 | EN-RAGE | P80511 |
| Fibroblast growth factor 19 | FGF-19 | O95750 |
| Fibroblast growth factor 21 | FGF-21 | Q9NSA1 |
| Fms-related tyrosine kinase 3 ligand | Flt3L | P49771 |
| Hepatocyte growth factor | HGF | P14210 |
| Interferon gamma | IFN-gamma | P01579 |
| Interleukin-10 receptor subunit beta | IL-10RB | Q08334 |
| Interleukin-12 subunit beta | IL-12B | P29460 |
| Interleukin-17C | IL-17C | Q9P0M4 |
| Interleukin-18 | IL-18 | Q14116 |
| Interleukin-18 receptor 1 | IL-18R1 | Q13478 |
| Interleukin-6 | IL-6 | P05231 |
| Interleukin-7 | IL-7 | P13232 |
| Interleukin-8 | IL-8 | P10145 |
| Interleukin-10 | IL10 | P22301 |
| Latency-associated peptide transforming growth factor beta-1 | LAP TGF-beta-1 | P01137 |
| Leukemia inhibitory factor receptor | LIF-R | P42702 |
| Monocyte chemotactic protein 1 | MCP-1 | P13500 |
| Monocyte chemotactic protein 2 | MCP-2 | P80075 |
| Monocyte chemotactic protein 3 | MCP-3 | P80098 |
| Monocyte chemotactic protein 4 | MCP-4 | Q99616 |
| Matrix metalloproteinase-1 | MMP-1 | P03956 |
| Matrix metalloproteinase-10 | MMP-10 | P09238 |
| Osteoprotegerin | OPG | O00300 |
| Oncostatin-M | OSM | P13725 |
| Programmed cell death ligand 1 | PD-L1 | Q9NZQ7 |
| Stem cell factor | SCF | P21583 |
| Sulfotransferase 1A1 | ST1A1 | P50225 |
| STAM-binding protein | STAMBP | O95630 |
| Transforming growth factor alpha | TGF-alpha | P01135 |
| Tumor necrosis factor | TNF | P01375 |
| TNF-beta | TNFB | P01374 |
| Tumor necrosis factor receptor superfamily member 9 | TNFRSF9 | Q07011 |
| Tumor necrosis factor ligand superfamily member 14 | TNFSF14 | O43557 |
| TNF-related apoptosis-inducing ligand | TRAIL | P50591 |
| TNF-related activation-induced cytokine | TRANCE | O14788 |
| Tumor necrosis factor (Ligand) superfamily, member 12 | TWEAK | O43508 |
| Urokinase-type plasminogen activator | uPA | P00749 |
| Vascular endothelial growth factor A | VEGF-A | P15692 |

**Table S3. List of 57 analysed metabolomic markers and their categories as classified by Nightingale Health.**

| Metabolite category | Metabolite (unit) |
| --- | --- |
| Lipoprotein particle size | Mean diameter for VLDL particles (nm) |
|  | Mean diameter for LDL particles (nm) |
|  | Mean diameter for HDL particles (nm) |
| Cholesterol | Serum total cholesterol (mmol/L) |
|  | Total cholesterol in VLDL (mmol/L) |
|  | Remnant cholesterol (mmol/L) |
|  | Total cholesterol in LDL (mmol/L) |
|  | Total cholesterol in HDL (mmol/L) |
|  | Total cholesterol in HDL2 (mmol/L) |
|  | Total cholesterol in HDL3 (mmol/L) |
|  | Esterified cholesterol (mmol/L) |
|  | Free cholesterol (mmol/L) |
| Glycerides and phospholipids | Serum total triglycerides (mmol/L) |
|  | Triglycerides in VLDL (mmol/L) |
|  | Triglycerides in LDL (mmol/L) |
|  | Triglycerides in HDL (mmol/L) |
|  | Total phosphoglycerides (mmol/L) |
|  | Ratio of triglycerides to phosphoglycerides |
|  | Phosphatidylcholine and other cholines (mmol/L) |
|  | Sphingomyelins (mmol/L) |
|  | Total cholines (mmol/L) |
| Apolipoproteins | Apolipoprotein A1 (g/L) |
|  | Apolipoprotein B (g/L) |
|  | Ratio of apolipoprotein B to apolipoprotein A1 |
| Fatty acids | Total fatty acids (mmol/L) |
|  | Degree of unsaturation |
|  | Docosahexaenoic acid (mmol/L) |
|  | Linoleic acid (mmol/L) |
|  | Omega-3 fatty acids (mmol/L) |
|  | Omega-6 fatty acids (mmol/L) |
|  | Polyunsaturated fatty acids (mmol/L) |
|  | Monounsaturated fatty acids (mmol/L) |
|  | Saturated fatty acids (mmol/L) |
| Fatty acid ratios | Ratio of docosahexaenoic acid to total fatty acids |
|  | Ratio of linoleic acid to total fatty acids |
|  | Ratio of omega-3 fatty acids to total fatty acids |
|  | Ratio of omega-6 fatty acids to total fatty acids |
|  | Ratio of polyunsaturated fatty acids to total fatty acids |
|  | Ratio of monounsaturated fatty acids to total fatty acids |
|  | Ratio of saturated fatty acids to total fatty acids |
| Glycolysis related metabolites | Glucose (mmol/L) |
|  | Lactate (mmol/L) |
|  | Citrate (mmol/L) |
| Amino acids | Alanine (mmol/L) |
|  | Glutamine (mmol/L) |
|  | Histidine (mmol/L) |
|  | Isoleucine (mmol/L) |
|  | Leucine (mmol/L) |
|  | Valine (mmol/L) |
|  | Phenylalanine (mmol/L) |
|  | Tyrosine (mmol/L) |
| Ketone bodies | Acetate (mmol/L) |
|  | Acetoacetate (mmol/L) |
|  | 3-hydroxybutyrate (mmol/L) |
| Fluid balance | Creatinine (mmol/L) |
|  | Albumin (signal area) |
| Inflammation | Glycoprotein acetyls (mmol/L) |

**Table S4. List of 28 analysed blood count and biochemistry markers.**

| Category | Marker (unit) | Assay kit (manufacturer) |
| --- | --- | --- |
| Full blood count | White blood cell count (10^9^/L) |  |
|  | Red blood cell count (10^12^/L) |  |
|  | Haemoglobin (g/L) |  |
|  | Haematocrit/packed cell volume (L/L) |  |
|  | Mean cell volume (fL) |  |
|  | Mean cell haemoglobin (pg) |  |
|  | Mean corpuscular haemoglobin concentration (g/L) |  |
|  | Platelet count (10^9^/L) |  |
|  | Neutrophils (10^9^/L) |  |
|  | Lymphocytes (10^9^/L) |  |
|  | Monocytes (10^9^/L) |  |
|  | Eosinophils (10^9^/L) |  |
|  | Basophils (10^9^/L) |  |
| Metabolic markers | Insulin (μU/mL) | Electrochemiluminescence immunoassay kit REF: 12017547 122 (Roche Diagnostics, Mannheim, Germany) |
|  | Glucose (mmol/L) | GLUC3 (Glucose HK) Cat. No. 04404483 190 (Roche Diagnostics, Mannheim, Germany) |
| Lipids | Total cholesterol (mmol/L) | CHOL2 (Cholesterol gen.2) Cat. No. 03039773 190 (Roche Diagnostics, Mannheim, Germany) |
|  | Triglycerides (mmol/L) | TRIGL (Triglycerides) Cat. No. 20767107 322 (Roche Diagnostics, Mannheim, Germany) |
|  | High-density lipoprotein (HDL) (mmol/L) | HDLC3 (HDL-Cholesterol plus 3rd generation) Cat. No. 04399803 190 (Roche Diagnostics, Mannheim, Germany) |
|  | Low-density lipoprotein (LDL) (mmol/L) | **Calculated using the Friedewald equation* |
|  | Very low-density lipoprotein (VLDL) (mmol/L) | **Calculated from triglycerides (VLDL = triglycerides / 2.19)* |
| Inflammation | C-reactive protein (CRP) (mg/L) | CRPHS (Cardiac C-Reactive Protein (Latex) High Sensitive) Cat. No. 04628918 190 (Roche Diagnostics, Mannheim, Germany) |
| Liver markers | Gamma-glutamyl transpeptide (GGT) (U/L) | GGT-2 (γ-Glutamyltransferase ver.2) System-ID: 0765988 (Roche Diagnostics, Mannheim, Germany) |
|  | Alanine aminotransferase (ALT) (U/L) | ALTLP (Alanine Aminotransferase acc. to IFCC with pyridoxal phosphate activation) System-ID: 07 6858 8 (Roche Diagnostics, Mannheim, Germany) |
|  | Aspartate aminotransferase (AST) (U/L) | ASTLP (Aspartate Aminotransferase acc. to IFCC with pyridoxal phosphate activation) System‑ID 07 6856 (Roche Diagnostics, Mannheim, Germany) |
|  | Procollagen-3 N-terminal peptide (P3NP) (μg/L) | UniQ PIIINP RIA Cat No. 68570 (Orion Diagnostica Oy, Espoo, Finland) |
|  | Hyaluronic acid (HA) (μg/L) | Corgenix, Colorado, USA |
| Derived | Homeostatic Model Assessment for Insulin Resistance (HOMA-IR) |  |
|  | AST/ALT |  |

**Table S5. Guidelines for Reporting on Latent Trajectory Studies (GRoLTS) Checklist.**

| GRoLTS checklist item | Yes/No | Comments |
| --- | --- | --- |
| 1. Is the metric of time used in the statistical model reported? | Yes |  |
| 2. Is information presented about the mean and variance of time within a wave? | N/A | Actual age in years was used as the time metric in the models rather than treating the data as time-structured. |
| 3a. Is the missing data mechanism reported? | Yes |  |
| 3b. Is a description provided of what variables are related to attrition/missing data? | N/A | The *lcmm* package allows individuals with missing data to be included in the models. |
| 3c. Is a description provided of how missing data in the analyses were dealt with? | Yes |  |
| 4. Is information about the distribution of the observed variables included? | Yes | See Table S1. |
| 5. Is the software mentioned? | Yes |  |
| 6a. Are alternative specifications of within-class heterogeneity considered (e.g., LCGA vs LGMM) and clearly documented? If not, was sufficient justification provided as to eliminate certain specifications from consideration? | Yes |  |
| 6b. Are alternative specifications of between-class differences in variance-covariance matrix structure considered and clearly documented? If not, was sufficient justification provided as to eliminate certain specifications from consideration? | Yes |  |
| 7. Are alternative shape/functional forms of the trajectories described? | Yes |  |
| 8. If covariates have been used, can analyses still be replicated? | N/A | Covariates were not included. |
| 9. Is information reported about the number of random start values and final iterations? | Yes |  |
| 10. Are the model comparison (and selection) tools described from a statistical perspective? | Yes |  |
| 11. Are the total number of fitted models reported, including a one-class solution? | Yes |  |
| 12. Are the number of cases per class reported for each model (absolute sample size, or proportion)? | Yes | See Table S6. |
| 13. If classification of cases in a trajectory is the goal, is entropy reported? | Yes | See Table S6. |
| 14a. Is a plot included with the estimated mean trajectories of the final solution? | Yes | See Figure 2. |
| 14b. Are plots included with the estimated mean trajectories for each model? | Yes | See Figure S2. |
| 14c. Is a plot included of the combination of estimated means of the final model and the observed individual trajectories split out for each latent class? | Yes | See Figure S3. |
| 15. Are characteristics of the final class solution numerically described (i.e., means, *SD*/*SE*, *n*, *CI*, etc.)? | Yes | See Tables 1 and S6. |
| 16. Are the syntax files available (either in the appendix, supplementary materials, or from the authors)? | Yes | See Methods S2. |

**Table S7. Characteristics of the ALSPAC subsample included in the biomarkers analyses, stratified by depression trajectories.**

|  | Low-stable  (n=1450) | Adolescent-limited  (n=369) | Adolescent-persistent  (n=137) | Adulthood-onset  (n=300) | *p* |
| --- | --- | --- | --- | --- | --- |
| Sex: Female | 739 (51.0%) | 261 (70.7%) | 109 (79.6%) | 190 (63.3%) | <0.001 |
| Ethnicity: Non-white | 44 (3.1%) | 6 (1.6%) | 6 (4.4%) | 11 (3.7%) | 0.281 |
| Maternal education | | | | | 0.059 |
| CSE or none | 100 (6.9%) | 23 (6.2%) | 19 (13.9%) | 19 (6.3%) |  |
| Vocational | 89 (6.1%) | 19 (5.1%) | 5 (3.6%) | 16 (5.3%) |  |
| O-level | 461 (31.8%) | 134 (36.3%) | 53 (38.7%) | 104 (34.7%) |  |
| A-level | 461 (31.8%) | 120 (32.5%) | 40 (29.2%) | 94 (31.3%) |  |
| Degree | 339 (23.4%) | 73 (19.8%) | 20 (14.6%) | 67 (22.3%) |  |
| Maternal occupational social class | | | | | 0.045 |
| I | 148 (10.2%) | 31 (8.4%) | 7 (5.1%) | 22 (7.3%) |  |
| II | 576 (39.7%) | 142 (38.5%) | 45 (32.8%) | 109 (36.3%) |  |
| III (non-manual) | 579 (39.9%) | 143 (38.8%) | 66 (48.2%) | 128 (42.7%) |  |
| III (manual) | 61 (4.2%) | 28 (7.6%) | 7 (5.1%) | 22 (7.3%) |  |
| IV or V^1^ | 86 (5.9%) | 25 (6.8%) | 12 (8.8%) | 19 (6.4%) |  |
| English IMD 2000 quintile | | | | | 0.271 |
| 1 – least deprived | 581 (40.7%) | 139 (38.2%) | 41 (30.1%) | 101 (34.1%) |  |
| 2 | 295 (20.7%) | 73 (20.1%) | 32 (23.5%) | 69 (23.3%) |  |
| 3 | 239 (16.8%) | 61 (16.8%) | 21 (15.4%) | 57 (19.3%) |  |
| 4 | 184 (12.9%) | 56 (15.4%) | 23 (16.9%) | 38 (12.8%) |  |
| 5 – most deprived | 127 (8.9%) | 35 (9.6%) | 19 (14.0%) | 31 (10.5%) |  |
| Family Adversity Index | 0 [0, 1] | 0 [0, 1] | 1 [0, 2] | 1 [0, 1] | 0.001 |
| BMI at age 10 | 17.43 [15.98, 19.53] | 17.36 [15.99, 19.41] | 18.23 [15.83, 21.02] | 17.30 [15.83, 19.34] | 0.200 |
| Number of SMFQ measurements | 8 [6, 10] | 9 [7, 10] | 8 [5, 9] | 9 [7, 10] | 0.001 |

^1^ Categories have been collapsed due to small cell counts.

Notes: Numbers presented as mean (SD), median [IQR], or n (%). Percentages are column percentages and computed based on the number of individuals with available data on each variable. Group comparisons were conducted using Chi-square tests for categorical variables and Kruskal-Wallis tests for non-normal continuous variables.

Abbreviations: CSE – Certificate of Secondary Education; O-level – Ordinary level; A-level – Advanced level; IMD – Index of Multiple Deprivation; SMFQ – Short Mood and Feelings Questionnaire

**Table S8. Associations of depression trajectories with cardiometabolic outcomes at 24 years and psychiatric outcomes at 24 and 28 years – unadjusted model**

|  |  | Adolescent-limited | | Adolescent-persistent | | Adulthood-onset |
| --- | --- | --- | --- | --- | --- | --- |
| Outcomes assessed at age 24 | n | Adjusted unstandardised regression coefficient (SE) | | | | |
| cIMT (continuous) | 1709 | -0.001 (0.003) | -0.009 (0.005) | | -0.002 (0.003) | |
| cfPWV (continuous) | 1860 | -0.139 (0.069) | **-0.285 (0.106)** | | -0.014 (0.077) | |
|  |  | Adjusted odds ratio (95% CI) | | | | |
| Smoking | 3085 | **1.58 (1.28-1.96)** | **2.44 (1.83-3.24)** | | **1.66 (1.31-2.08)** | |
| AUDIT-C score ≥ 5 | 3055 | 0.89 (0.73-1.09) | **0.57 (0.43-0.75)** | | **0.78 (0.63-0.97)** | |
| BMI ≥ 30kg/m^2^ | 3082 | 1.35 (1.01-1.80) | **2.59 (1.82-3.63)** | | 1.23 (0.88-1.69) | |
| Elevated waist circumference | 3054 | **1.32 (1.07-1.63)** | **1.81 (1.35-2.42)** | | 1.11 (0.87-1.40) | |
| Triglycerides ≥1.7mmol/L | 2539 | 1.14 (0.75-1.70) | 0.92 (0.46-1.67) | | 1.18 (0.75-1.80) | |
| HDL <1.0mmol/L | 2539 | 1.04 (0.76-1.40) | 1.46 (0.97-2.15) | | 1.37 (1.01-1.85) | |
| SBP ≥ 130mmHg | 3109 | 0.66 (0.48-0.91) | **0.60 (0.36-0.95)** | | 0.82 (0.58-1.13) | |
| DBP ≥ 85mmHg | 3109 | 1.34 (0.66-2.53) | 1.54 (0.58-3.45) | | 1.26 (0.56-2.53) | |
| Fasting glucose ≥5.6mmol/L | 2539 | **0.67 (0.51-0.87)** | 0.87 (0.59-1.24) | | 0.88 (0.57-1.15) | |
| Metabolic syndrome | 3120 | 0.96 (0..61-1.45) | 1.28 (0.72-2.14) | | 1.35 (1.01-1.80) | |
| ICD-10 depressive episode | 3085 | **3.81 (2.64-5.47)** | **24.08 (16.85-34.68)** | | **8.03 (5.74-11.30)** | |
| ICD-10 GAD | 3074 | **3.15 (2.19-4.51)** | **17.42 (12.25-24.90)** | | **4.80 (3.37-6.82)** | |
| Outcomes assessed at age 28 |  |  |  | |  | |
| Prescribed antidepressants in past 5y | 3361 | **3.07 (2.39-3.94)** | **7.70 (5.79-10.24)** | | **4.96 (3.87-6.34)** | |
| Prescribed anxiolytics in past 5y | 3410 | **2.21 (1.25-3.77)** | **5.98 (3.57-9.90)** | | **4.08 (2.50-6.60)** | |

Notes: Effect estimates presented are unstandardised regression coefficients for continuous outcomes and odds ratios (95% confidence intervals) for binary outcomes. The reference group for all analyses is the low-stable trajectory. Text in bold indicates evidence for the association after FDR correction of p-values.

Metabolic syndrome was defined based on the 2009 consensus definition from the International Diabetes Federation and the American Heart Association/National Heart, Lung, and Blood Institute, i.e. the presence of any three of the following five risk factors: elevated waist circumference (≥94cm for white men, ≥90cm for non-white men, ≥80cm for women); elevated triglycerides (≥1.7mmol/L), reduced high-density lipoprotein-cholesterol (<1.0mmol/L), elevated blood pressure (systolic ≥130mmHg or diastolic ≥85mmHg), and elevated fasting glucose (≥5.6mmol/L).

Abbreviations: AUDIT-C – Alcohol Use Disorders Identification Test for Consumption; BMI – body mass index; cfPWV – carotid-femoral pulse wave velocity; cIMT – carotid intima-media thickness; DBP – diastolic blood pressure; GAD – generalised anxiety disorder; HDL – high-density lipoprotein; ICD-10 – International Classification of Diseases 10^th^ Revision; SBP – systolic blood pressure
